# Retrospective cohort study extracting coexisting background breast-lesion features from stage I-III invasive breast cancer

**DOI:** 10.64898/2026.05.19.26353633

**Authors:** Ryan Jak Yang Lim, Phyu Nitar, Kah Weng Lau, Lester Chee Hao Leong, Geok Hoon Lim, Veronique Kiak Mien Tan, Benita Kiat Tee Tan, Ern Yu Tan, Serene Si Ning Goh, Mikael Hartman, Fuh Yong Wong, Jingmei Li, Joint Breast Cancer Registry

**Affiliations:** Genome Institute of Singapore (GIS), Agency for Science, Technology and Research (A*STAR), Singapore, Singapore; Department of Cancer Informatics, National Cancer Centre Singapore, Singapore, Singapore; Department of Pathology, National University Hospital, National University Health System, Singapore, Singapore; Department of Diagnostic Radiology, Khoo Teck Puat Hospital, Singapore, Singapore; Breast Department, KK Women’s and Children’s Hospital, Singapore, Singapore; Division of Surgery and Surgical Oncology, National Cancer Centre Singapore, Singapore Health Services, Singapore, Singapore; Department of Breast Surgery, Singapore General Hospital, Singapore Health Services, Singapore, Singapore; SingHealth Duke-NUS Breast Centre, Singapore, Singapore; Department of Surgery, Sengkang General Hospital, Singapore, Singapore; Department of General Surgery, Tan Tock Seng Hospital, Singapore, Singapore; Lee Kong Chian School of Medicine, Singapore, Singapore; Institute of Molecular and Cell Biology (IMCB), Agency for Science, Technology and Research (A*STAR), Singapore, Singapore; Department of Surgery, Yong Loo Lin School of Medicine, National University of Singapore, Singapore, Singapore; Department of Surgery, National University Health System, Singapore, Singapore; Division of Radiation Oncology, National Cancer Centre Singapore, Singapore Health Services, Singapore, Singapore; National Cancer Centre Singapore, Singapore Health Services, Singapore, Singapore

**Keywords:** Natural language processing (NLP), pathology, free-text, data extraction, structured variables, benign breast diseases, histological features

## Abstract

**Background:** Background breast features are frequently noted in pathology reports alongside invasive breast cancer but rarely factor into prognosis or treatment decisions. Their relationship to tumor characteristics and patient outcomes remains incompletely characterised.

**Methods:** We conducted a retrospective cohort study of 7,603 patients with Stage I–III invasive breast cancer (diagnosed 1991–2022, age <80 years) from the Joint Breast Cancer Registry in Singapore. Natural language processing (NLP) was applied to 9,754 free-text pathology reports to extract co-existing background breast features, with accuracy validated by dual-reviewer assessment of 200 reports. Unsupervised hierarchical clustering grouped extracted features into three categories. Associations with tumor characteristics were assessed by multinomial logistic regression, and ten-year overall survival by Cox proportional hazards models (median follow-up 9.6 years; 620 deaths).

**Results:** Here we show that NLP-based extraction of background breast features from routine pathology reports achieves an accuracy of over 90% across features. Lobular neoplasia and benign proliferative changes are associated with less aggressive tumor characteristics, whereas early neoplastic and papillary lesions are more prevalent in HER2-enriched and luminal B tumor subtypes. Benign proliferative changes are associated with better survival in age- and year-adjusted models (hazard ratio 0.91, 95% CI 0.86–0.97), but this association is attenuated after adjustment for stage and subtype.

**Conclusions:** NLP-enabled extraction of background breast features from pathology text is feasible at scale. These features reflect tumor biology but do not independently add prognostic information beyond established clinical variables.

**PLAIN LANGUAGE SUMMARY:** When a patient is diagnosed with breast cancer, the pathologist examining the tissue sample may also observe other changes in the surrounding breast, such as benign growths, cysts, or early precancerous lesions. These incidental findings are often recorded in free-text notes within pathology reports but are rarely studied in large numbers because collecting them by hand is impractical. We used a computer program that reads medical text (natural language processing) to extract these background findings automatically from over 9,700 pathology reports collected across 30 years. We found that certain background changes are linked to specific tumor types, but they do not independently predict which patients will do better or worse once tumor stage and type are considered. This work shows that valuable information in pathology reports can be automatically extracted at scale, opening new opportunities to study the tissue environment in which breast cancers develop.

## INTRODUCTION

Most breast cancer research focuses on tumor molecular, genetic, and histologic characteristics because of their direct relevance to prognosis and treatment. In contrast, coexisting background breast features in pathology reports, such as benign, atypical, or proliferative lesions, are often considered incidental and receive less attention. Moreover, these features are inconsistently and unsystematically reported in pathology reports, often appearing only in free-text notes. This lack of standardized reporting, combined with challenges in data extraction, has limited large-scale research into the relevance of coexisting breast features in invasive breast cancer (IBC). While ductal carcinoma in situ (DCIS), a recognized non-invasive precursor to IBC, has been studied, the prognostic significance of other benign or high-risk features noted in pathology reports remains unclear (1).

With the increasing digitization of health records, natural language processing (NLP) enables large-scale extraction of clinically relevant data from pathology narratives (2-4). This study aims to characterize the tumor characteristics of Stage I–III IBC patients with coexisting breast features extracted via NLP, and to explore their associations with prognosis. We show that NLP-based extraction of background breast features from routine pathology reports is feasible and achieves >90% accuracy across all features examined. Background lesions cluster into three biologically coherent groups and are associated with distinct tumor characteristics. However, they do not independently predict overall survival after accounting for stage and molecular subtype.

## METHODS

### Participants

The Joint Breast Cancer Registry (JBCR) is a hospital-based registry that compiles comprehensive data on breast cancer patients who have been diagnosed or treated within institutions under the SingHealth Cluster in Singapore. Participating sites include Singapore General Hospital (SGH), National Cancer Centre Singapore (NCCS), KK Women’s and Children’s Hospital (KKH), Changi General Hospital (CGH), and Sengkang General Hospital (SKH) (5). The registry captures a wide range of information, including patient demographics, tumor characteristics (such as date of diagnosis, histology, stage, size, nodal involvement, and hormone receptor status), treatment details, adverse drug reactions, and survival outcomes.

The causes and dates of death are retrieved from the National Registry of Births and Deaths by utilizing each patient’s National Registration Identity Card (NRIC) number as a unique identifier to link clinical data (6). All live births, deaths, and stillbirths in Singapore must be reported within a designated timeframe. Mandatory reporting requirements minimize the risk of loss to follow-up. A dashboard within the SingHealth enterprise data warehouse (electronic Health INTelligence systems; eHINTs) allows for real-time data retrieval and visualization (5).

Informed consent approval from the SingHealth Centralised Institutional Review Board is waived for JBCR (CIRB reference: 2019/2419). This study is approved by A*STAR Institutional Review Board (2024-105).

### Extraction and processing of free-text pathology data

Pathology reports were obtained from the SingHealth-Synapxe Electronic Health Intelligence System (eHints), a centralized data repository that integrates clinical data across multiple healthcare institutions within the SingHealth network. All reports were de-identified by the registry’s data management team and a trusted third party within the SingHealth cluster.

We used natural language processing (NLP, OpenAI API, “GPT-4-turbo”, temperature=0) to systematically process and extract structured information from unstructured free-text pathology reports. Five distinct prompt templates were developed to perform targeted extractions:

Prompt 1: Identify whether the pathology report described breast tissue and extract a supporting quote when applicable.

Prompt 2: Extract the specimen type and determine the presence of malignancy. Accepted specimen types included excision, vacuum-assisted biopsy (VAB), core needle biopsy (CNB), fine-needle aspiration (FNA), incisional biopsy, skin punch biopsy, sentinel lymph node biopsy, or “unknown.”

Prompt 3: Identify the presence of specific pre-invasive breast lesions (i.e. ductal carcinoma in situ (DCIS), lobular carcinoma in situ (LCIS), atypical ductal hyperplasia (ADH), atypical lobular hyperplasia (ALH)) and determine the laterality (left, right, bilateral, or unknown).

Prompt 4: Detect benign proliferative breast lesions, including usual ductal hyperplasia (UDH), sclerosing adenosis, radial scar/complex sclerosing lesion, intraductal papilloma, and fibroadenoma.

Prompt 5: Detect benign non-proliferative breast lesions such as cysts, apocrine metaplasia, columnar cell change, fibrocystic change, calcifications, and flat epithelial atypia (FEA).

Atypical features were extracted as named diagnostic entities (ADH via Prompt 3; FEA via Prompt 5) rather than via a generic keyword search for “atypia” or “atypical”, to avoid false positives from non-specific uses of these terms in pathology language. Cases in which both intraductal papilloma and ADH were co-reported (consistent with an atypical papilloma) were captured through independent extraction by Prompts 3 and 4.

To assess the accuracy of NLP-based extraction, we manually validated a stratified random sample of 200 pathology reports (∼5% of the dataset). Sampling was stratified using an 8-bin design based on the number of benign features detected by the API (0, 1, 2, or ≥3) and the character length of the report (<4000 vs. ≥4000 characters). Fifteen reports were sampled per bin (n=120), and the remaining 80 were allocated proportionally, based on the distribution of cases across bins in the full dataset. Two study team members (a data analyst [RJYL] and a pathologist [KWL]) independently reviewed each report to confirm the presence or absence of the histological features studied.

Unsupervised hierarchical clustering was conducted in R to identify patterns in the presence or absence of breast features across reports. A pairwise Pearson correlation matrix was computed based on binary feature vectors (presence=1, absence=0). To transform similarity into a dissimilarity measure, the distance matrix was defined as 1 minus the Pearson correlation coefficient. Clustering was performed using the hclust() function with the Ward.D2 method, which minimizes the total within-cluster variance, allowing for compact and interpretable clustering of samples based on shared breast feature profiles.

### Statistics and reproducibility

Multinomial logistic regression (“nnet” package in R) was performed to evaluate associations between specific breast lesions and tumor clinicopathological features. Reference categories were defined as Stage I, ER-positive, PR-positive, tumor size <2□cm, well-differentiated grade, node-negative status, and Luminal A proxy subtype. To account for multiple hypothesis testing, we applied the Benjamini-Hochberg procedure to control the false discovery rate (FDR). Adjusted p-values were calculated using the p.adjust() function in R.

We used Cox proportional hazards regression models to evaluate the association between coexisting breast lesions and overall survival at 10 years. We limited our analysis to Stage I–III invasive breast cancer in patients under 80 years of age to focus on a clinically and biologically homogeneous group most likely to benefit from standard curative-intent treatments. We also excluded patients aged 80 years and older who often have multiple comorbidities and face higher mortality from competing conditions. The outcome variable was time from diagnosis to death or censoring at 10 years. Patients alive beyond 10 years were censored at that time point. Cox models were fitted using the coxph() function from the survival R package. The proportional hazards assumption in Cox regression was formally assessed using the cox.zph() function. Sensitivity analyses were conducted by specimen type (excision, CNB), disease stage (Stage I, Stage II/III), and calendar period (before 2010, 2010 onwards, to account for the implementation of AJCC 7th⍰edition staging and broader use of HER2-directed therapies).

Logistic regression was used to estimate odds ratios (ORs) for the presence of any benign breast feature, while Poisson regression was used to estimate rate ratios (RRs) for the total number of benign features. Stage I served as the reference group in both models.

To assess reproducibility, NLP extraction was validated against independent dual-reviewer assessment (one data analyst and one pathologist) in a stratified random sample of 200 reports. All analyses were conducted in R (v4.5.0) and the full source code is publicly available (see Code Availability).

## RESULTS

### Study cohort

**Supplementary Figure 1** shows the derivation of the analytical dataset. We processed 90,417 free-text histopathology reports. Of these, 54,292 were confirmed as breast tissue; 26,145 showed malignancy. We retained 19,196 reports that involved excision (n=8,221), CNB (n=9,378), VAB (n=682), or FNA (n=915). After removing duplicates, the cleaned pathology set comprising 15,774 reports was obtained. From the clinical registry, 29,092 entries were deduplicated to 29,058 unique unilateral breast cancer patients. Merging the two datasets yielded 15,210 reports from 11,281 patients. Limiting to pathology reports dated within 3 months before to 6 months after diagnosis gave 12,756 reports (10,092 patients). After restricting to Stage I–III and age <80, the final dataset included 9,754 reports from 7,603 patients (diagnosed 1991-2022). The median age at diagnosis is 56.0 years (IQR: 47.0–64.0). Median BMI of 23.9 kg/m^2^ (IQR: 21.3–27.2) (**Table 1**). Most were diagnosed from 2010 onwards (62%), and the majority were of Chinese ethnicity (77%).

**Table 1.** Patient characteristics at breast cancer diagnosis.

| Characteristic | All<br>(n=7,603) | CNB<br>(n=4,571) | Excision<br>(n=3,988) | CNB (n=4,571) |  |  | Excision (n = 3,988) |  |  |
| --- | --- | --- | --- | --- | --- | --- | --- | --- | --- |
|  |  |  |  | Stage I<br>(n=1,371) | Stage II<br>(n=2,111) | Stage III<br>(n=1,089) | Stage I<br>(n=1,604) | Stage II<br>(n=1,634) | Stage III<br>(n=750) |
| Age, years,<br>median (IQR) | 56.0<br>(47.0–64.0) | 57.0<br>(48.0–65.0) | 55.0<br>(47.0–64.0) | 58.0<br>(50.0–66.0) | 57.0<br>(48.0–66.0) | 56.0<br>(48.0–64.0) | 55.0<br>(47.0–63.0) | 55.0<br>(46.0–64.0) | 55.0<br>(47.0–64.0) |
| Body mass index, kg/m2,<br>median (IQR) | 23.9<br>(21.3–27.2) | 24.1<br>(21.5–27.5) | 23.9<br>(21.2–27.2) | 23.4<br>(21.2–26.6) | 24.4<br>(21.7–27.7) | 24.3<br>(21.5–28.0) | 23.5<br>(21.1–26.4) | 24.2<br>(21.4–27.5) | 24.1<br>(21.2–27.8) |
| Year of diagnosis |  |  |  |  |  |  |  |  |  |
| Pre-2002 | 616 (8) | 73 (2) | 330 (8) | 5 (0) | 36 (1) | 32 (1) | 82 (2) | 203 (5) | 45 (1) |
| 2002-2009 | 2,237 (29) | 1,135 (25) | 1,426 (36) | 315 (7) | 513 (11) | 307 (7) | 539 (14) | 595 (15) | 292 (7) |
| 2010 and later | 4,750 (62) | 3,363 (74) | 2,232 (56) | 1,051 (23) | 1,562 (34) | 750 (16) | 983 (25) | 836 (21) | 413 (10) |
| Ethnicity (n, %) |  |  |  |  |  |  |  |  |  |
| Chinese | 5829 (77) | 3,466 (76) | 3,086 (77) | 1,151 (25) | 1,596 (35) | 719 (16) | 1,339 (34) | 1,242 (31) | 505 (13) |
| Indian | 387 (5) | 222 (5) | 214 (5) | 45 (1) | 113 (2) | 64 (1) | 62 (2) | 101 (3) | 51 (1) |
| Malay | 717 (9) | 490 (11) | 328 (8) | 83 (2) | 228 (5) | 179 (4) | 75 (2) | 154 (4) | 99 (2) |
| Others | 670 (9) | 393 (9) | 360 (9) | 92 (2) | 174 (4) | 127 (3) | 128 (3) | 137 (3) | 95 (2) |
| Marital status (n, %) |  |  |  |  |  |  |  |  |  |
| Married | 5,162 (68) | 3,058 (67) | 2,741 (69) | 907 (20) | 1,416 (31) | 735 (16) | 1,109 (28) | 1,113 (28) | 519 (13) |
| Single/Divorced/Separated/Widowed | 1,569 (21) | 1,044 (23) | 791 (20) | 282 (6) | 470 (10) | 292 (6) | 289 (7) | 327 (8) | 175 (4) |
| Others | 872 (11) | 469 (10) | 456 (11) | 182 (4) | 225 (5) | 62 (1) | 206 (5) | 194 (5) | 56 (1) |
| Number of children (n, %) |  |  |  |  |  |  |  |  |  |
| None | 482 (6) | 300 (7) | 223 (6) | 96 (2) | 132 (3) | 72 (2) | 75 (2) | 86 (2) | 62 (2) |
| 1 | 352 (5) | 211 (5) | 184 (5) | 58 (1) | 97 (2) | 56 (1) | 71 (2) | 66 (2) | 47 (1) |
| 2 | 847 (11) | 544 (12) | 409 (10) | 178 (4) | 255 (6) | 111 (2) | 177 (4) | 154 (4) | 78 (2) |
| ≥3 | 855 (11) | 536 (12) | 446 (11) | 132 (3) | 277 (6) | 127 (3) | 134 (3) | 199 (5) | 113 (3) |
| Unknown | 5,067 (67) | 2,980 (65) | 2,726 (68) | 907 (20) | 1,350 (30) | 723 (16) | 1,147 (29) | 1,129 (28) | 450 (11) |
| Menopausal status (n, %) |  |  |  |  |  |  |  |  |  |
| Post | 3,821 (50) | 2,456 (54) | 2,003 (50) | 793 (17) | 1,095 (24) | 568 (12) | 850 (21) | 762 (19) | 391 (10) |
| Pre or peri | 1,853 (24) | 1,058 (23) | 1,024 (26) | 309 (7) | 494 (11) | 255 (6) | 421 (11) | 399 (10) | 204 (5) |
| Unknown | 1,929 (25) | 1,057 (23) | 961 (24) | 269 (6) | 522 (11) | 266 (6) | 333 (8) | 473 (12) | 155 (4) |
| Family history of breast cancer (n, %) |  |  |  |  |  |  |  |  |  |
| Yes | 1,013 (13) | 651 (14) | 516 (13) | 224 (5) | 305 (7) | 122 (3) | 240 (6) | 197 (5) | 79 (2) |
| No | 4,790 (63) | 2,968 (65) | 2,557 (64) | 892 (20) | 1,326 (29) | 750 (16) | 1,025 (26) | 991 (25) | 541 (14) |
| Unknown | 1,800 (24) | 952 (21) | 915 (23) | 255 (6) | 480 (11) | 217 (5) | 339 (9) | 446 (11) | 130 (3) |

### NLP extraction performance

The two most common specimen types were CNB (n=4,571) and excisions (n=3,988) (**Supplementary Figure 1, Table 1**). The median time from CNB to diagnosis was 0 days, while the median time between diagnosis and excision was 21 days (IQR: 5 - 35) (**Supplementary Figure 2**). A total of 1,398 patients had pathology reports from both CNB and excision procedures. Agreement between CNB and excision samples for identifying B3 features was generally low, with Cohen’s kappa ranging from −0.003 to 0.336 (**Supplementary Table 1**). Notably, the proportion of breast features identified in excision reports consistently exceeded those observed in CNB reports across all lesion subtypes (**Supplementary Table 1**). The presence or absence of breast features from the excision dataset (n=3,988) was carried forward for further analyses.

The data analyst (RJYL) manually reviewed 200 NLP-generated structured reports and found 51 (25.5%) with at least one disagreement across all breast features studied. KWL independently reviewed the same 200 cases and disagreed with RJYL on 3 of those 51 instances with discrepancies. In all cases, the pathologist’s opinion (KWL) was considered the gold standard. When considered individually, we observed that the breast features had an overall accuracy of over 90% (**Table 2**). Sensitivity ranged from 0.75 to 1.00 and specificity from 0.93 to 1.00 which suggested strong agreement between NLP-derived labels and human annotation.

**Table 2.** Performance of natural language processing (NLP) in extracting histological features, based on manual review of 200 pathology reports. Metrics reported include sensitivity, specificity, positive predictive value (PPV), negative predictive value (NPV), and overall accuracy, benchmarked against manual verification as the reference standard. True positives (TP), false positives (FP), false negatives (FN), and true negatives (TN) are also provided.

|  | Sensitivity | Specificity | PPV | NPV | Accuracy | TP | FP | FN | TN |
| --- | --- | --- | --- | --- | --- | --- | --- | --- | --- |
| Lobular neoplasia |  |  |  |  |  |  |  |  |  |
| Lobular carcinoma in situ (LCIS) | 0.818 | 1 | 1 | 0.990 | 0.990 | 9 | 0 | 2 | 189 |
| Atypical lobular hyperplasia (ALH) | 0.750 | 1 | 1 | 0.995 | 0.995 | 3 | 0 | 1 | 196 |
| Benign or non-atypical proliferative breast changes |  |  |  |  |  |  |  |  |  |
| Fibroadenoma | 0.929 | 0.930 | 0.500 | 0.994 | 0.930 | 13 | 13 | 1 | 173 |
| Calcification | 1 | 0.954 | 0.875 | 1 | 0.965 | 49 | 7 | 0 | 144 |
| Cyst | 0.926 | 0.994 | 0.962 | 0.989 | 0.985 | 25 | 1 | 2 | 172 |
| Apocrine metaplasia | 1 | 0.989 | 0.905 | 1 | 0.990 | 19 | 2 | 0 | 179 |
| Columnar cell change | 1 | 0.995 | 0.933 | 1 | 0.995 | 14 | 1 | 0 | 185 |
| Usual ductal hyperplasia (UDH) | 0.929 | 1 | 1 | 0.989 | 0.990 | 26 | 0 | 2 | 172 |
| Sclerosing adenosis | 0.929 | 0.978 | 0.765 | 0.995 | 0.975 | 13 | 4 | 1 | 182 |
| Fibrocystic change | 1 | 0.987 | 0.957 | 1 | 0.990 | 45 | 2 | 0 | 153 |
| Early neoplastic, papillary and complex sclerosing lesions |  |  |  |  |  |  |  |  |  |
| Ductal carcinoma in situ (DCIS) | 0.906 | 0.932 | 0.958 | 0.850 | 0.915 | 115 | 5 | 12 | 68 |
| Atypical ductal hyperplasia (ADH) | 0.875 | 1 | 1 | 0.995 | 0.995 | 7 | 0 | 1 | 192 |
| Flat epithelial atypia (FEA) | 1 | 1 | 1 | 1 | 1 | 2 | 0 | 0 | 198 |
| Radial scar or complex sclerosing lesion | 1 | 0.985 | 0.667 | 1 | 0.985 | 6 | 3 | 0 | 191 |
| Intraductal papilloma | 1 | 0.995 | 0.909 | 1 | 0.995 | 10 | 1 | 0 | 189 |

### Unsupervised clustering of breast features

Hierarchical clustering of breast features revealed three distinct groups: (1) lobular neoplasia (LCIS, ALH), (2) benign or non-atypical proliferative breast changes with minimal to low associated cancer risk (i.e., fibroadenoma, calcification, cyst, apocrine metaplasia, columnar cell change, UDH, sclerosing adenosis, fibrocystic change), and (3) early neoplastic (DCIS, ADH, FEA), papillary and complex sclerosing lesions (**Supplementary Figure 3**).

### Associations with tumor characteristics

**Table 3** shows the associations between breast features and tumor characteristics in 3,988 breast cancer cases with reports from excision procedures. Lobular neoplasia (LCIS and ALH) was generally associated with less aggressive tumor features, including lower odds of advanced stage (Stage II, OR_with reference to Stage I_ 0.74 [0.57-0.97] and Stage III, OR_with reference to Stage I_ 0.68 [0.48-0.97]) hormone receptor negativity (OR_ER-negative_ 0.37 [0.25-0.54] and OR_PR-negative_ 0.59 [0.45-0.78]), poor differentiation (OR_with reference to well-differentiated_ 0.34 [0.23-0.49]), and aggressive proxy subtypes (lower odds for Luminal B, HER2-enriched and triple-negative subtypes after multiple testing correction). However, LCIS, but not ALH, was associated with increased odds of larger tumor size (≥5 cm, OR 1.75 [1.13-2.71]).

**Table 3.**
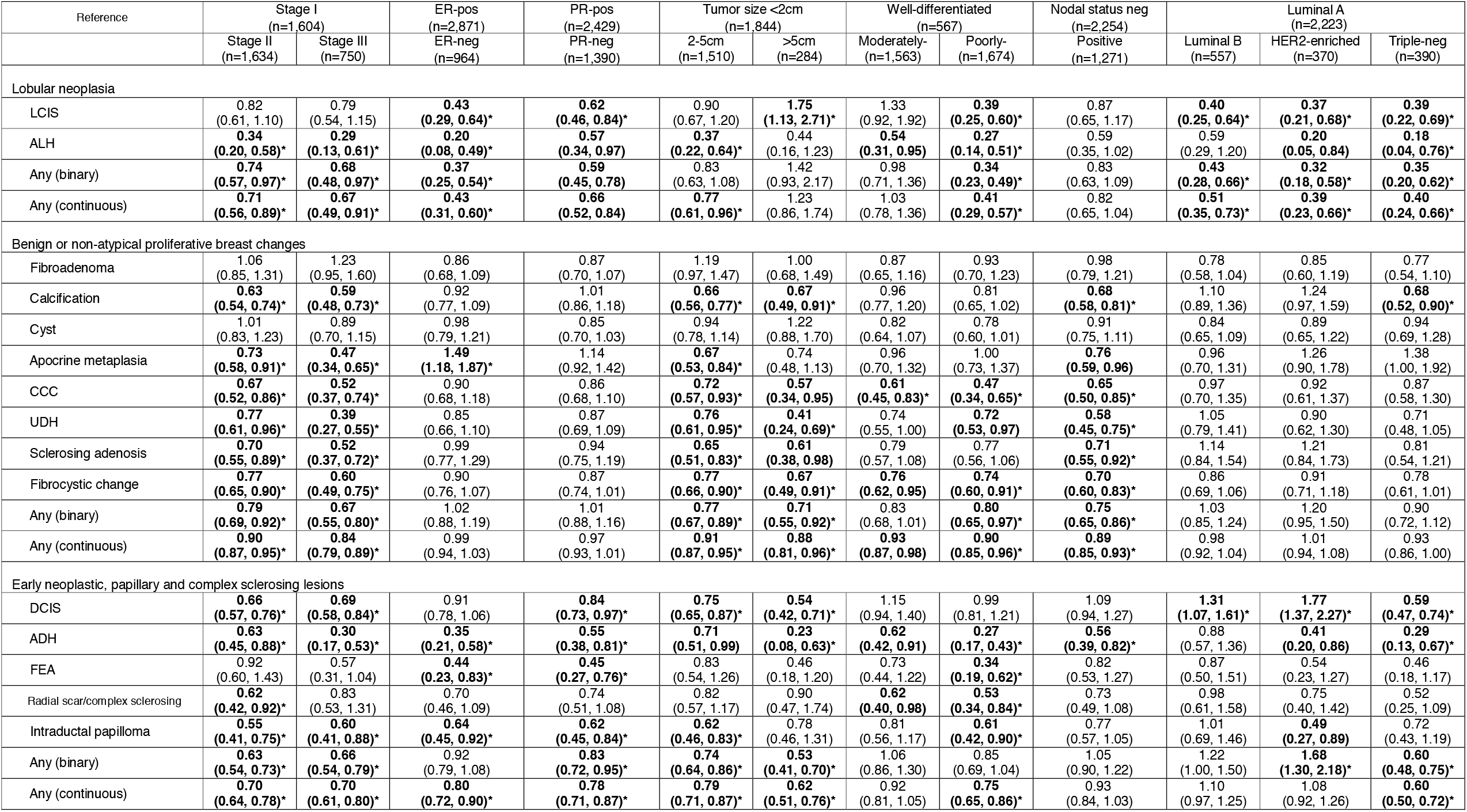
Association between histological breast features and tumor characteristics in 3,988 breast cancer cases with reports from excision procedures. Odds ratios (ORs) and 95% confidence intervals (CIs) are presented from multinomial logistic regression models evaluating the association between breast features and tumor characteristics at diagnosis. Each OR reflects the odds of having a specific tumor characteristic versus a reference category, given the presence of a feature. All models are adjusted for potential confounders, including age at diagnosis, year of diagnosis, ethnicity, family history of breast cancer, menopausal status, and parity. All tests were two-sided. Statistically significant associations (p < 0.05) are indicated in bold. Multiple comparisons were controlled using the Benjamini–Hochberg procedure (false discovery rate adjustment); associations remaining significant after correction are denoted by *. LCIS: Lobular carcinoma in situ; ALH: Atypical lobular hyperplasia; CCC: Columnar cell change; UDH: Usual ductal hyperplasia; DCIS: Ductal carcinoma in situ; ADH: Atypical ductal hyperplasia; FEA: Flat epithelial atypia

Similarly, benign and non-atypical proliferative breast changes were generally associated with a lower likelihood of advanced stage (Stage II, OR_with reference to Stage I_ 0.79 [0.69-0.92] and Stage III, OR_with reference to Stage I_ 0.67 [0.55-0.80]), larger tumor size (2-5 cm, OR_with reference to <2 cm_ 0.77 [0.67-0.89] and >5 cm, OR_with reference to <2 cm_ 0.71 [0.55-0.92]), poorly differentiated tumors (OR_with reference to well-differentiated_ 0.80 [0.65-0.97]), and positive node involvement (OR 0.89 [0.85-0.93]), with the exception of fibroadenoma and cyst. Apocrine metaplasia was associated with higher odds of ER-negative tumors (OR 1.49 [1.18–1.87]).

The presence of any early neoplastic, papillary and complex sclerosing lesions (binary) was significantly lower in more advanced tumor stages (Stage II, OR_with reference to Stage I_ 0.63 [0.54-0.73] and Stage III, OR_with reference to Stage I_ 0.66 [0.54-0.79]). Similarly, tumors measuring 2–5 cm and >5 cm were significantly less likely to have early lesions than those <2 cm (OR 0.74 [0.64-0.86] and OR 0.53 [0.41-0.70]). With respect to hormone receptor status, PR-negative tumors had a significantly lower prevalence of these features than PR-positive tumors (OR 0.83 [0.72-0.95]). Among molecular subtypes, triple-negative breast cancers showed a reduced likelihood of harboring such features (OR 0.60 [0.48–0.75]), while HER2-enriched subtypes had significantly higher odds compared to Luminal A (OR 1.68 [1.30–2.18]). DCIS was associated with higher odds of Luminal B (OR 1.31 [1.07-1.61]) and HER2-enriched subtypes (OR 1.77 [1.37-2.27]).

A sensitivity analysis of the CNB and calendar period subsets showed that generally, point estimates were still protective, however, fewer associations were statistically significant (**Supplementary Tables 2-4**).

### Ten-year overall survival

In 3,988 breast cancer patients (620 deaths, median follow-up time: 9.6 years), any benign or non-atypical proliferative breast changes (binary) variable was associated with 10-year overall survival (log-rank p=0.00089) (**Figure 1**). The any benign or non-atypical proliferative breast changes (continuous) variable showed a significant positive association with survival (HR 0.91 [0.86-0.97]) before factoring for tumor characteristics. In particular, calcifications were associated with improved survival in partially adjusted models (HR 0.73 [0.59-0.89]). However, no breast feature was significantly associated with survival after further adjusting for stage and proxy subtype (**Table 4**). Results of sensitivity analyses by subgroups based on specimen type, calendar period and disease stage are shown in **Supplementary Tables 5-10**.

**Figure 1.**
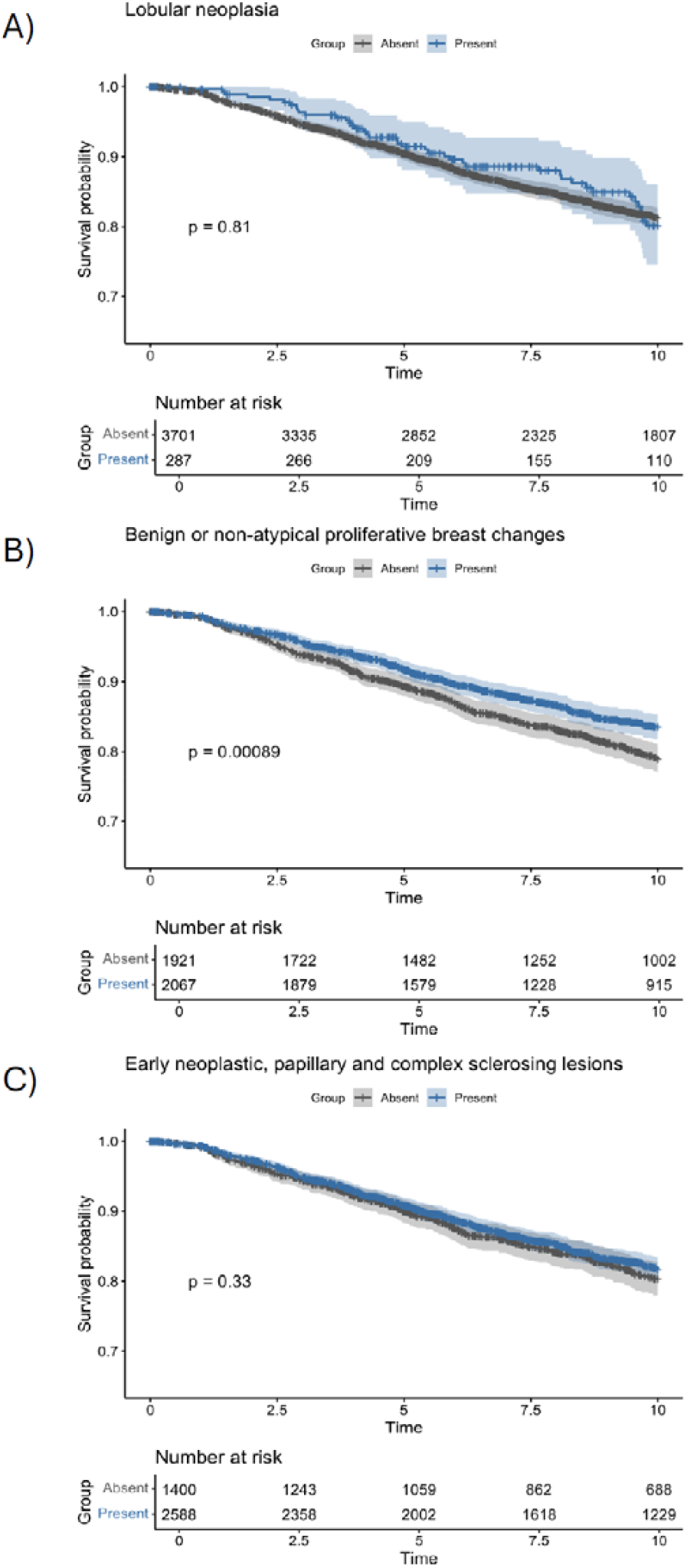
Kaplan–Meier curves showing 10-year overall survival. Comparisons of presence or absence of A) lobular neoplasia, B) benign or non-atypical proliferative breast changes, and C) early neoplastic, papillary and complex sclerosing lesions, among 3,988 breast cancer cases with reports from excision procedures. Shaded bands represent 95% confidence intervals for the survival estimates. Log-rank tests were used for group comparisons (two-sided). Tick marks indicate censored observations. The source data for this figure is in Supplementary Data 1.

**Table 4.** Association between coexisting histological breast features and 10-year overall survival in 3,988 breast cancer patients (620 events). Hazard ratios (HRs) with 95% confidence intervals (CIs) are presented from Cox proportional hazards models evaluating the impact of coexisting histological breast features on 10-year overall survival among breast cancer patients. All tests were two-sided. Statistically significant associations (p < 0.05) are indicated in bold. Multiple comparisons were controlled using the Benjamini–Hochberg procedure (false discovery rate adjustment); associations remaining significant after correction are denoted by *. a) Adjusted for age at diagnosis and year of diagnosis. b) Model one further adjusted for menstruation status, ethnicity, family history of cancer and parity. c) Model two further adjusted for tumor characteristics: stage and subtype.

|  | Number of patients | Number of events | Model 1<sup>a</sup><br>HR (95% CI) | Model 2<sup>b</sup><br>HR (95% CI) | Model 3<sup>c</sup><br>HR (95% CI) |
| --- | --- | --- | --- | --- | --- |
| **Lobular neoplasia** |  |  |  |  |  |
| Lobular carcinoma in situ (LCIS) | 246 (6.2%) | 36 (14.6%) | 1.02 (0.73, 1.43) | 1.02 (0.72, 1.43) | 1.09 (0.78, 1.54) |
| Atypical lobular hyperplasia (ALH) | 80 (2.0%) | 8 (10.0%) | 0.76 (0.38, 1.52) | 0.77 (0.38, 1.54) | 0.95 (0.47, 1.92) |
| Any (binary) | 287 (7.2%) | 41 (14.3%) | 1.02 (0.74, 1.40) | 1.02 (0.74, 1.41) | 1.12 (0.81, 1.55) |
| Any (continuous) | - | - | 0.96 (0.73, 1.27) | 0.97 (0.73, 1.27) | 1.05 (0.80, 1.39) |
| **Benign or non-atypical proliferative breast changes** |  |  |  |  |  |
| Fibroadenoma | 491 (12.3%) | 67 (13.6%) | 0.96 (0.74, 1.24) | 0.93 (0.72, 1.20) | 0.93 (0.72, 1.21) |
| Calcification | 1,020 (25.6%) | 116 (11.4%) | **0.73 (0.60, 0.90)\*** | **0.73 (0.59, 0.89)\*** | 0.82 (0.67, 1.01) |
| Cyst | 626 (15.7%) | 74 (11.8%) | 0.83 (0.65, 1.06) | 0.79 (0.62, 1.01) | 0.81 (0.63, 1.03) |
| Apocrine metaplasia | 423 (10.6%) | 48 (11.3%) | 0.78 (0.58, 1.05) | 0.78 (0.58, 1.05) | 0.91 (0.67, 1.22) |
| Columnar cell change | 348 (8.7%) | 36 (10.3%) | 0.77 (0.55, 1.09) | 0.79 (0.56, 1.11) | 0.87 (0.62, 1.22) |
| Usual ductal hyperplasia (UDH) | 413 (10.4%) | 45 (10.9%) | 0.74 (0.55, 1.01) | **0.73 (0.54, 1.00)** | 0.91 (0.67, 1.24) |
| Sclerosing adenosis | 373 (9.4%) | 46 (12.3%) | 0.91 (0.67, 1.23) | 0.93 (0.68, 1.25) | 1.11 (0.82, 1.51) |
| Fibrocystic change | 1042 (26.1%) | 128 (12.3%) | **0.81 (0.67, 0.99)** | 0.84 (0.69, 1.02) | 0.95 (0.78, 1.15) |
| Any (binary) | 2067 (51.8%) | 275 (13.3%) | **0.81 (0.69, 0.95)** | **0.80 (0.68, 0.94)** | 0.88 (0.75, 1.04) |
| Any (continuous) | - | - | **0.92 (0.86, 0.97)\*** | **0.91 (0.86, 0.97)\*** | 0.96 (0.90, 1.01) |
| **Early neoplastic, papillary and complex sclerosing lesions** |  |  |  |  |  |
| Ductal carcinoma in situ (DCIS) | 2,449 (61.4%) | 367 (15.0%) | 0.93 (0.79, 1.10) | 0.93 (0.79, 1.09) | 1.04 (0.89, 1.23) |
| Atypical ductal hyperplasia (ADH) | 170 (4.3%) | 14 (8.2%) | **0.53 (0.31, 0.91)** | **0.54 (0.32, 0.92)** | 0.69 (0.41, 1.18) |
| Flat epithelial atypia (FEA) | 108 (2.7%) | 8 (7.4%) | 0.60 (0.30, 1.21) | 0.62 (0.30, 1.26) | 0.71 (0.35, 1.44) |
| Radial scar or complex sclerosing lesion | 148 (3.7%) | 17 (11.5%) | 0.80 (0.49, 1.29) | 0.79 (0.49, 1.28) | 0.77 (0.48, 1.26) |
| Intraductal papilloma | 240 (6.0%) | 41 (17.1%) | 1.18 (0.86, 1.63) | 1.19 (0.86, 1.64) | **1.39 (1.01, 1.93)** |
| Any (binary) | 2588 (64.9%) | 393 (15.2%) | 0.97 (0.82, 1.15) | 0.97 (0.82, 1.14) | 1.09 (0.92, 1.29) |
| Any (continuous) | - | - | 0.92 (0.81, 1.03) | 0.91 (0.81, 1.03) | 1.01 (0.90, 1.14) |

In the analysis of tumor stage associations with breast features, both Stage II and Stage III were associated with a lower number of breast features compared to Stage I (logistic regression p<5.28E-04, Poisson regression p<2.61E-21) (**Supplementary Table 11**).

## DISCUSSION

This analysis reveals several interesting findings. Firstly, NLP accurately extracted structured variables from pathology reports, with sensitivity and specificity above 90%. Secondly, unsupervised clustering of breast features revealed three distinct groups: A) lobular neoplasia, B) benign or non-atypical proliferative breast changes, and C) early neoplastic, papillary and complex sclerosing lesions. Thirdly, the presence of any breast features was generally linked to less aggressive tumor characteristics, with benign or non-atypical proliferative breast changes associated with better prognosis. Despite these associations, all breast features studied were not significantly linked to long-term survival after adjusting for tumor characteristics.

Recent work shows that large language models like ChatGPT can accurately extract structured data from pathology reports without task-specific training (7, 8). In lung cancer and pediatric osteosarcoma datasets, ChatGPT-3.5 achieved up to 98–100% accuracy for key pathological features, outperforming traditional NLP methods (9). Performance depended on prompt design and was reproducible over time. In another study, a ChatGPT-based Streamlit app structured 33 breast cancer pathology reports with 99.6% accuracy that significantly reducing processing time (10).

To our knowledge, no prior study has applied unsupervised clustering to text-extracted coexisting breast features from pathology reports. The close pathological and clinical relationship between LCIS and ALH makes their clustering as lobular neoplasia biologically and clinically expected (11). The second cluster of breast features may share common hormonal and microenvironmental influences. Fibroadenomas, cysts, apocrine metaplasia, and UDH are estrogen-responsive and arise from the terminal duct lobular unit, making their co-occurrence anatomically and biologically plausible (12). Columnar cell change and sclerosing adenosis have also been previously described to be frequently observed together (13). The emergence of these clusters demonstrates the potential of NLP not only to extract information but also to uncover latent biological patterns that may be overlooked in conventional analysis. Most prior work using NLP in pathology relies on supervised learning, where models are trained to predict predefined labels such as cancer subtype, grade, or biomarker status, based on annotated training data (14). While powerful, supervised approaches are inherently constrained by existing clinical classifications and may miss novel groupings or previously unrecognized feature relationships. In contrast, unsupervised learning, as applied here, does not require labelled outcomes and instead allows the data to self-organize based on intrinsic similarity. This exploratory framework is particularly valuable in histopathology, where coexisting lesions may interact in complex, non-linear ways that are not well-captured by current taxonomies.

The traditional linear model of breast carcinogenesis by Wellings et al. describes progression from normal epithelium through FEA, ADH, DCIS, to invasive cancer (15-17). Interestingly, in our study, lesions such as radial scars or complex sclerosing lesions and intraductal papillomas are often observed with FEA, ADH, and DCIS. Their co-occurrence may not reflect a strict linear sequence but rather shared tissue-level processes, such as epithelial proliferation, stromal remodeling, or hormonal influences, that create a microenvironment conducive to early neoplastic change. This observed heterogeneity highlights the complexity of breast carcinogenesis and suggests that multiple, biologically distinct pathways may lead to malignancy. Further research is warranted to elucidate the temporal and mechanistic relationships between these coexisting features.

The association between benign or early neoplastic lesions and less aggressive tumor subtypes may reflect distinct biological pathways leading to indolent cancers or increased clinical surveillance enabling earlier detection. For instance, advances in molecular subtype research suggest that low-grade luminal A cancers may follow a stepwise progression from atypia, providing time for precursor lesions to develop (18). In contrast, high-grade subtypes such as TNBC and HER2+ cancers may arise de novo, without identifiable precursors, indicating a fundamentally different tumor trajectory (19). Calcifications, while not true histopathologic tissue features, are routinely reported because they correlate strongly with underlying breast pathology and are key radiologic markers for follow-up and detection (20). Their presence may therefore act as a surrogate for increased diagnostic scrutiny, indirectly contributing to the observed association between background features and earlier-stage or less aggressive disease. Additionally, certain histologic subtypes, such as DCIS or columnar cell change, are more likely to present with calcifications, further linking this feature to indolent disease pathways (21, 22).

Coexisting breast features are often underreported, inconsistently described, or omitted if not clinically prioritized. Reporting may also vary by stage, where satisfaction of search and anchoring are common cognitive biases (23). In early-stage disease, background tissue is more often detailed to guide surveillance, while late-stage reports focus on tumor and biomarker status, overlooking benign features. Indeed, when we compared breast feature counts across cancer stages, we found earlier-stage cases had more features recorded, consistent with reduced reporting depth in advanced disease. It should be noted that stage-dependent reporting bias may confound clinical interpretation by making certain breast features appear more prognostically relevant than they are (i.e. a reflection of more documentation in early-stage cases rather than true biological effects). Also, in advanced stage disease, the tumor may be sizeable and extensive. As such, there may be limited amount of “normal background tissue” to examine.

This study uses a large, multi-institutional hospital-based registry with rich clinical and pathological data to explore the co-occurrence of breast features. A key strength lies in the use of NLP to extract structured histological information from unstructured pathology reports. The NLP-generated dataset was validated using stratified random sampling and dual human review to ensure high accuracy and reliability.

However, the study is limited by its retrospective design, and the pathology reports may not have reported all the co-existing breast lesions. The fraction of histologically identifiable background features that pathologists chose not to document (especially high-stage or biopsy-only specimens) remains unknown and cannot be estimated from pathology text alone. A prospective slide re-review study in a smaller representative cohort would be needed to quantify this reporting gap and to assess whether the associations we describe reflect true biological co-occurrence or documentation biases. There may also be variability in pathology report formats over time. The prognosis of the patients may have been affected by other factors such as the treatment which they received, though the management of breast cancer patients were usually discussed in a multidisciplinary meeting and decided predominantly based on the cancer characteristics.

Several limitations of the NLP pipeline warrant acknowledgement. DCIS extraction showed the lowest sensitivity in validation (0.906), with 12 false negatives from 200 sampled reports. The prompts did not explicitly specify how the model should handle hedged pathology language such as “consistent with” or “suspicious for” and we did not conduct a structured analysis of failure cases. For example, whether misclassifications clustered around ambiguous formulations, specific report types, or specimen characteristics. Without such an error analysis, the primary causes of misclassification remain uncertain. Prompt refinement informed by systematic error review, as well as benchmarking against newer proprietary or open-source models, would be valuable directions for future work. We used GPT-4-turbo (temperature = 0) to ensure deterministic, reproducible extraction, but more recent models may offer improved accuracy or reduced operational cost.

The primary contribution of this study is methodological. We demonstrated that structured extraction of background features from unstructured pathology text is feasible and scalable. While our findings are not immediately practice-changing, they help place breast cancer in a broader pathological context. Further research is needed to confirm their clinical value. Meanwhile, our study highlights the potential value of more comprehensive pathology reporting and supports future risk stratification or treatment tailoring strategies based on routinely observed but currently underutilized histologic features.

## Joint Breast Cancer Registry

Ahmed Syed Salahuddin^1^, Alcantara Veronica Siton^2^, Beh Sok Yuen^3^, Chan Johan^3^, Chan Junjie Jack^3^, Chay Wen Yee^3^, Cheng Chee Leong^1^, Cherylin Tan Ruiling^1^, Chew Ming Long Melvin^3^, Chew Sui Tjien Lita^3^, Chong Jun Hua^4^, Chua Eu Tiong^3^, Chua Gail Wan Ying^3^, Chua Hui Wen^5^, Chua Ji Guang Bernard^3^, Dent Rebecca^3^, Ewe See Hooi^4^, Fok Wai Yee Rose^3^, Gudi Mihir^2^, Hamzah Julie Liana Bte^1^, Hing Jun Xian^7^, Ho Shihan Bryan^3^, Hwee Lin Wee^8^, Iqbal Jabed^1^, Jain Amit^3^, Koh Wee Yao^9^, Koo Si-Lin^3^, Kwan Xin Yi Clara Anne^5^, Lee Chee Meng^5^, Lee Chuan Yaw^3^, Lee Han Yi^3^, Lee Hui Ting Stefanie^1^, Lee Jie Xin Joycelyn^3^, Lee Su Xin Ghislaine^3^, Leong Qi Hui Faith^5^, Lim Faye^3^, Lim Hsuen Elaine^3^, Lim Li Hoon^3^, Lim Sheng Hao Joshua^1^, Ma Jun^3^, Mohd Ishak Hanis Mariyah Binte^3^, Mok Chi Wei^7^, Nei Wen Long^3^, Ng Choon Ta^4^, Ng Raymond^3^, Ng Wee Loon^3^, Ngeow Yuen Yie Joanne^3^, Pang Siyan Jinnie^2^, Phyu Nitar^3^, Preetha Madhukumar^3^, Shih Vivianne^3^, Sim Pei Yin Dayna^1^, Sim Yirong^3^, Tan Boon Fei^3^, Tan Hong Qi^3^, Tan Jing Ying Tira^3^, Tan Kuan Rui Lloyd^3^, Tan Ngiap Chuan^10^, Tan Qing Ting^2^, Tan Ser Huey Janice^3^, Tan Si Ying^1^, Tan Su-Ming^7^, Tan Ying Ching^1^, Tan Ying Cong Ryan Shea^3^, Tan Yongcheng Benjamin^1^, Tan Zhi Chien Joshua^3^, Tay Kwang Yong Timothy^1^, Teo Wei Ling^3^, Tey Min Li^3^, Wong Mabel^3^, Wong Su Lin Jill^3^, Yan Zhiyan^2^, Yang Shi-Hui Christina^1^, Yap Yoon Sim^3^, Yeo Khung Keong^4^, Yeo Ming Chert Richard^3^, Yeong Poh Sheng Joe^1^, Yit Ling Fung Nelson^3^, Yong Wei Sean^3,^ & Zhang Zewen^3^

^1^Singapore General Hospital, Singapore, Singapore. ^2^KK Women’s and Children’s Hospital, Singapore, Singapore. ^3^National Cancer Centre Singapore, Singapore, Singapore. ^4^National Heart Centre Singapore, Singapore, Singapore. ^5^Sengkang General Hospital, Singapore, Singapore. ^6^Duke-NUS, Singapore, Singapore. ^7^Changi General Hospital, Singapore, Singapore. ^8^NUS-Saw Swee Hock School of Public Health, Singapore, Singapore. ^9^National University Hospital, Singapore, Singapore. ^10^SingHealth Polyclinic - Head Office, Singapore, Singapore.

## DECLARATIONS

### Ethics approval and consent to participate

The JBCR is approved by the SingHealth Centralized Institutional Review Board (CIRB 2019/2419). As the study utilizes de-identified, registry-based data, the requirement for informed consent was waived. This study is approved by A*STAR IRB (2024-105).

### Consent for publication

Not applicable.

### Data Availability

Individual-level patient data from the Joint Breast Cancer Registry (JBCR) are held by the National Cancer Centre Singapore (NCCS) and cannot be made publicly available owing to regulatory and data governance requirements. De-identified data supporting the conclusions of this study are available upon reasonable request, subject to institutional data access agreements. Requests should be directed to the JBCR data access committee via https://www.nccs.com.sg/research-innovation/data-on-request; a response will be provided within 15 working days of receipt.

### Code Availability

The full source code, including all NLP prompt templates and statistical analysis scripts, is publicly available at https://github.com/ryan-limjy/Histopathology-paper.

### Conflict of interest and financial disclosures

The authors declare that they have no competing interests

### Funding

This study is funded by the Agency for Science, Technology and Research (A*STAR). RL is a recipient of an A*STAR Research Internship Award (ARIA).

### Authors’ contributions

J.L., S.G., E.Y.T., and F.Y.W. conceptualised the study and designed the methodology. P.N. and F.Y.W. performed data acquisition and curation. R.J.Y.L. conducted all data analyses. K.W.L. participated in manual validation of NLP extraction as the pathologist reviewer. L.C.H.L., G.H.L., V.K.M.T., B.K.T.T., E.Y.T., S.G., M.H., and F.Y.W. contributed to patient data through the Joint Breast Cancer Registry and provided clinical expertise in the interpretation of results. R.J.Y.L. and J.L. drafted the manuscript. All authors interpreted the results, reviewed, revised, and approved the final version of the manuscript.

### AI declaration

The original content and research presented in this manuscript were fully authored by us. The assistance of Grammarly and ChatGPT was used to exclusively enhance language clarity and readability (Jun, 2025). All scientific content, interpretations, and conclusions remain the sole responsibility of the authors.

## Acknowledgements

Not applicable.

## Notes

### Competing Interest Statement

The authors have declared no competing interest.

