## Supplementary Information for "Retrospective cohort study extracting coexisting background breast-lesion features from stage I-III invasive breast cancer"

**Supplementary Table 1.** Comparison of breast features detected by core needle biopsy (green) and excision (orange) procedures. The table below shows the concordance between lesion presence on core needle biopsy (CNB) and subsequent surgical excision in 1,398 patients with both records. Cohen's kappa (κ) is reported to assess agreement beyond chance. κ values closer to 1 indicate stronger agreement.

**Supplementary Table 2.** Association between breast features and tumor characteristics in 3,164 breast cancer cases with reports from core needle biopsies. Odds ratios (ORs) and 95% confidence intervals (CIs) are presented from multinomial logistic regression models evaluating the association between breast features and tumor characteristics at diagnosis. Each OR reflects the odds of having a specific tumor characteristic versus a reference category, given the presence of a breast feature. All models are adjusted for potential confounders, including age at diagnosis, year of diagnosis, ethnicity, family history of breast cancer, menopausal status, and parity. All tests were two-sided. Statistically significant associations ($p < 0.05$) are indicated in bold. Multiple comparisons were controlled using the Benjamini–Hochberg procedure (false discovery rate adjustment); associations remaining significant after correction are denoted by *. LCIS: Lobular carcinoma in situ; ALH: Atypical lobular hyperplasia; CCC: Columnar cell change; UDH: Usual ductal hyperplasia; DCIS: Ductal carcinoma in situ; ADH: Atypical ductal hyperplasia; FEA: Flat epithelial atypia

**Supplementary Table 3.** Association between breast features and tumor characteristics in 1,756 breast cancer cases with reports from excision procedures, diagnosed before 2010. Odds ratios (ORs) and 95% confidence intervals (CIs) are presented from multinomial logistic regression models evaluating the association between breast features and tumor characteristics at diagnosis. Each OR reflects the odds of having a specific tumor characteristic versus a reference category, given the presence of a feature. All models are adjusted for potential confounders, including age at diagnosis, year of diagnosis, ethnicity, family history of breast cancer, menopausal status, and parity. All tests were two-sided. Statistically significant associations ($p < 0.05$) are indicated in bold. Multiple comparisons were controlled using the Benjamini–Hochberg procedure (false discovery rate adjustment); associations remaining significant after correction are denoted by *. LCIS: Lobular carcinoma in situ; ALH: Atypical lobular hyperplasia; CCC: Columnar cell change; UDH: Usual ductal hyperplasia; DCIS: Ductal carcinoma in situ; ADH: Atypical ductal hyperplasia; FEA: Flat epithelial atypia

**Supplementary Table 4.** Association between breast features and tumor characteristics in 2,232 breast cancer cases with reports from excision procedures, diagnosed 2010 and after. Odds ratios (ORs) and 95% confidence intervals (CIs) are presented from multinomial logistic regression models evaluating the association between breast features and tumor characteristics at diagnosis. Each OR reflects the odds of having a specific tumor characteristic versus a reference category, given the presence of a feature. All models are adjusted for potential confounders, including age at diagnosis, year of diagnosis, ethnicity, family history of breast cancer, menopausal status, and parity. All tests were two-sided. Statistically significant associations ($p < 0.05$) are indicated in bold. Multiple comparisons were controlled using the Benjamini–Hochberg procedure (false discovery rate adjustment); associations remaining significant after correction are denoted by *. LCIS: Lobular carcinoma in situ; ALH: Atypical lobular hyperplasia; CCC: Columnar cell change; UDH: Usual ductal hyperplasia; DCIS: Ductal carcinoma in situ; ADH: Atypical ductal hyperplasia; FEA: Flat epithelial atypia

**Supplementary Table 5.** Association between coexisting features and 10-year overall survival in 3,164 breast cancer patients with CNB reports (613 events). Hazard ratios (HRs) with 95% confidence intervals (CIs) are presented from Cox proportional hazards models evaluating the impact of coexisting breast features on 10-year overall survival among breast cancer patients. All tests were two-sided. Statistically significant associations ($p < 0.05$) are indicated in bold. Multiple comparisons were controlled using the Benjamini–Hochberg procedure (false discovery rate adjustment); associations remaining significant after correction are denoted by *. a) Adjusted for age at diagnosis and year of diagnosis. b) Model one further adjusted for menstruation status, ethnicity, family history of cancer and parity. c) Model two further adjusted for tumour characteristics: stage and subtype.

**Supplementary Table 6.** Association between coexisting features and 10-year overall survival in 1,756 breast cancer cases with reports from excision procedures, diagnosed before 2010. Hazard ratios (HRs) with 95% confidence intervals (CIs) are presented from Cox proportional hazards models evaluating the impact of coexisting breast features on 10-year overall survival among breast cancer patients. All tests were two-sided. Statistically significant associations ($p < 0.05$) are indicated in bold. Multiple comparisons were controlled using the Benjamini–Hochberg procedure (false discovery rate adjustment); associations remaining significant after correction are denoted by *. a) Adjusted for age at diagnosis and year of diagnosis. b) Model one further adjusted for menstruation status, ethnicity, family history of cancer and parity. c) Model two further adjusted for tumour characteristics: stage and subtype.

**Supplementary Table 7.** Association between coexisting features and 10-year overall survival in 2,232 breast cancer cases with reports from excision procedures, diagnosed 2010 and after. Hazard ratios (HRs) with 95% confidence intervals (CIs) are presented from Cox proportional hazards models evaluating the impact of coexisting breast features on 10-year overall survival among breast cancer patients. All tests were two-sided. Statistically significant associations ($p < 0.05$) are indicated in bold. Multiple comparisons were controlled using the Benjamini–Hochberg procedure (false discovery rate adjustment); associations remaining significant after correction are denoted by *. a) Adjusted for age at diagnosis and year of diagnosis. b) Model one further adjusted for menstruation status, ethnicity, family history of cancer and parity. c) Model two further adjusted for tumour characteristics: stage and subtype.

**Supplementary Table 8.** Association between coexisting features and 10-year overall survival in 1,604 breast cancer cases with reports from excision procedures diagnosed with Stage I breast cancer. Hazard ratios (HRs) with 95% confidence intervals (CIs) are presented from Cox proportional hazards models evaluating the impact of coexisting breast features on 10-year overall survival among breast cancer patients. All tests were two-sided. Statistically significant associations ($p < 0.05$) are indicated in bold. Multiple comparisons were controlled using the Benjamini–Hochberg procedure (false discovery rate adjustment); associations remaining significant after correction are denoted by *. a) Adjusted for age at diagnosis and year of diagnosis. b) Model one further adjusted for menstruation status, ethnicity, family history of cancer and parity. c) Model two further adjusted for tumour characteristics: subtype.

**Supplementary Table 9.** Association between coexisting features and 10-year overall survival in 1,634 breast cancer cases with reports from excision procedures diagnosed with Stage II breast cancer. Hazard ratios (HRs) with 95% confidence intervals (CIs) are presented from Cox proportional hazards models evaluating the impact of coexisting breast features on 10-year overall survival among breast cancer patients. All tests were two-sided. Statistically significant associations ($p < 0.05$) are indicated in bold. Multiple comparisons were controlled using the Benjamini–Hochberg procedure (false discovery rate adjustment); associations remaining significant after correction are denoted by *. a) Adjusted for age at diagnosis and year of diagnosis. b) Model one further adjusted for menstruation status, ethnicity, family history of cancer and parity. c) Model two further adjusted for tumour characteristics: subtype.

**Supplementary Table 10.** Association between coexisting features and 10-year overall survival in 750 breast cancer cases with reports from excision procedures diagnosed with Stage III breast cancer. Hazard ratios (HRs) with 95% confidence intervals (CIs) are presented from Cox proportional hazards models evaluating the impact of coexisting breast features on 10-year overall survival among breast cancer patients. All tests were two-sided. Statistically significant associations ($p < 0.05$) are indicated in bold. Multiple comparisons were controlled using the Benjamini–Hochberg procedure (false discovery rate adjustment); associations remaining significant after correction are denoted by *. a) Adjusted for age at diagnosis and year of diagnosis. b) Model one further adjusted for menstruation status, ethnicity, family history of cancer and parity. c) Model two further adjusted for tumour characteristics: subtype.

**Supplementary Table 11.** Association between tumor stage and presence/number of breast features, using 3,988 breast cancer cases with reports from excision procedures. All tests were two-sided. No multiple comparison adjustment was applied as analyses were pre-specified and limited in number.

**Supplementary Figure 1.** Flowchart of how analytical datasets were derived.

**Supplementary Figure 2.** Distribution of procedure date relative to diagnosis date.

**Supplementary Figure 3.** Cluster membership of breast features based on Pearson correlation hierarchical clustering for 3,988 records from excisions.

**Supplementary Table 1.** Comparison of breast features detected by core needle biopsy (green) and excision (orange) procedures. The table below shows the concordance between lesion presence on core needle biopsy (CNB) and subsequent surgical excision in 1,398 patients with both records. Cohen's kappa (κ) is reported to assess agreement beyond chance. κ values closer to 1 indicate stronger agreement.

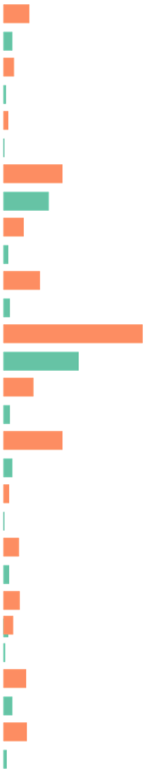

| | Present (n, %) | CNB (+) & Excision (+) | CNB (+) & Excision (−) | CNB (−) & Excision (+) | CNB (−) & Excision (−) | Cohen's κ |
|---|---|---|---|---|---|---|
| Apocrine metaplasia | | 22 (1.6%) | 35 (2.5%) | 142 (10.2%) | 1,199 (85.8%) | 0.148 |
| Atypical ductal hyperplasia (ADH) | | 2 (0.1%) | 14 (1.0%) | 65 (4.6%) | 1,317 (94.2%) | 0.03 |
| Atypical lobular hyperplasia (ALH) | | 1 (0.1%) | 8 (0.6%) | 30 (2.1%) | 1,359 (97.2%) | 0.04 |
| Calcification | | 134 (9.6%) | 157 (11.2%) | 243 (17.4%) | 864 (61.8%) | 0.217 |
| Columnar cell change | | 3 (0.2%) | 32 (2.3%) | 127 (9.1%) | 1,236 (88.4%) | -0.003 |
| Cyst | | 16 (1.1%) | 29 (2.1%) | 216 (15.5%) | 1,137 (81.3%) | 0.065 |
| DCIS | | 386 (27.6%) | 95 (6.8%) | 497 (35.6%) | 420 (30.0%) | 0.217 |
| Fibroadenoma | | 18 (1.3%) | 24 (1.7%) | 175 (12.5%) | 1,181 (84.5%) | 0.109 |
| Fibrocystic change | | 21 (1.5%) | 37 (2.6%) | 355 (25.4%) | 985 (70.5%) | 0.027 |
| Flat epithelial atypia (FEA) | | 1 (0.1%) | 7 (0.5%) | 39 (2.8%) | 1,351 (96.6%) | 0.032 |
| Intraductal papilloma | | 8 (0.6%) | 29 (2.1%) | 91 (6.5%) | 1,270 (90.8%) | 0.082 |
| LCIS | | 24 (1.7%) | 7 (0.5%) | 79 (5.7%) | 1,288 (92.1%) | 0.336 |
| Radial scar or complex sclerosing lesion | | 6 (0.4%) | 7 (0.5%) | 58 (4.1%) | 1,327 (94.9%) | 0.143 |
| Sclerosing adenosis | | 9 (0.6%) | 51 (3.6%) | 135 (9.7%) | 1,203 (86.1%) | 0.029 |
| Usual ductal hyperplasia | | 5 (0.4%) | 19 (1.4%) | 146 (10.4%) | 1,228 (87.8%) | 0.028 |

**Supplementary Table 2.** Association between breast features and tumor characteristics in 3,164 breast cancer cases with reports from core needle biopsies. Odds ratios (ORs) and 95% confidence intervals (CIs) are presented from multinomial logistic regression models evaluating the association between breast features and tumor characteristics at diagnosis. Each OR reflects the odds of having a specific tumor characteristic versus a reference category, given the presence of a breast feature. All models are adjusted for potential confounders, including age at diagnosis, year of diagnosis, ethnicity, family history of breast cancer, menopausal status, and parity. All tests were two-sided. Statistically significant associations ($p < 0.05$) are indicated in bold. Multiple comparisons were controlled using the Benjamini–Hochberg procedure (false discovery rate adjustment); associations remaining significant after correction are denoted by *. LCIS: Lobular carcinoma in situ; ALH: Atypical lobular hyperplasia; CCC: Columnar cell change; UDH: Usual ductal hyperplasia; DCIS: Ductal carcinoma in situ; ADH: Atypical ductal hyperplasia; FEA: Flat epithelial atypia

| Reference | Stage I (n=801) | | ER-pos (n=2,357) | PR-pos (n=2,005) | Tumor size <2cm (n=1,115) | | Well-differentiated (n=340) | | Nodal status neg (n=1,441) | Luminal A (n=1,824) | | |
|---|---|---|---|---|---|---|---|---|---|---|---|---|
| | Stage II (n=1,537) | Stage III (n=826) | ER-neg (n=746) | PR-neg (n=1,096) | 2-5cm (n=1,395) | >5cm (n=259) | Moderately- (n=1,174) | Poorly- (n=1,376) | Positive (n=1,125) | Luminal B (n=559) | HER2-enriched (n=291) | Triple-neg (n=335) |
| Lobular neoplasia | | | | | | | | | | | | |
| LCIS | **0.51 (0.28, 0.91)*** | 0.58 (0.29, 1.15) | **0.30 (0.12, 0.76)*** | 0.71 (0.40, 1.28) | 0.96 (0.52, 1.77) | 1.59 (0.66, 3.83) | 1.03 (0.48, 2.18) | **0.33 (0.14, 0.78)*** | 1.07 (0.59, 1.93) | 0.47 (0.21, 1.06) | **0.13 (0.02, 0.97)** | **0.22 (0.05, 0.92)** |
| ALH | 0.36 (0.10, 1.31) | 0.17 (0.02, 1.44) | 0.31 (0.04, 2.47) | 0.19 (0.02, 1.47) | 0.43 (0.11, 1.74) | 1.56 (0.31, 7.90) | 0.20 (0.03, 1.19) | 0.32 (0.07, 1.45) | 0.53 (0.14, 2.11) | 0.39 (0.05, 3.13) | - | 0.61 (0.08, 4.94) |
| Any (binary) | **0.49 (0.28, 0.86)*** | 0.55 (0.28, 1.06) | **0.33 (0.14, 0.77)*** | 0.67 (0.38, 1.18) | 0.87 (0.49, 1.56) | 1.81 (0.82, 4.00) | 0.87 (0.43, 1.74) | **0.33 (0.15, 0.73)*** | 1.09 (0.63, 1.90) | **0.43 (0.19, 0.96)** | **0.12 (0.02, 0.87)** | **0.31 (0.09, 0.99)** |
| Any (continuous) | **0.52 (0.32, 0.86)*** | **0.54 (0.29, 0.99)** | **0.34 (0.15, 0.77)*** | 0.64 (0.38, 1.09) | 0.86 (0.51, 1.43) | 1.46 (0.72, 2.97) | 0.85 (0.46, 1.57) | **0.37 (0.18, 0.75)*** | 0.96 (0.58, 1.59) | 0.50 (0.24, 1.02) | **0.13 (0.02, 0.92)** | **0.32 (0.10, 0.99)** |
| Benign or non-atypical proliferative breast changes | | | | | | | | | | | | |
| Fibroadenoma | 1.3 (0.84, 2.10) | 0.57 (0.30, 1.09) | 0.98 (0.62, 1.56) | 0.86 (0.56, 1.30) | 1.14 (0.76, 1.73) | 0.32 (0.10, 1.03) | 0.93 (0.50, 1.73) | 0.73 (0.39, 1.36) | 0.93 (0.61, 1.43) | 1.12 (0.68, 1.84) | 1.29 (0.69, 2.38) | 0.68 (0.32, 1.43) |
| Calcification | **0.68 (0.55, 0.84)*** | **0.71 (0.56, 0.91)*** | **0.65 (0.51, 0.81)*** | **0.77 (0.63, 0.93)*** | 0.83 (0.68, 1.01) | **0.61 (0.41, 0.89)*** | 1.01 (0.76, 1.35) | **0.63 (0.47, 0.85)*** | 1.17 (0.96, 1.43) | 1.23 (0.98, 1.55) | 1.10 (0.81, 1.50) | **0.40 (0.27, 0.59)*** |
| Cyst | **0.5 (0.33, 0.85)*** | **0.36 (0.18, 0.69)*** | 0.73 (0.42, 1.28) | 0.69 (0.42, 1.12) | 0.77 (0.47, 1.25) | 0.85 (0.37, 1.96) | 0.70 (0.38, 1.27) | **0.33 (0.17, 0.64)*** | **0.42 (0.24, 0.73)*** | 0.66 (0.35, 1.24) | 0.50 (0.20, 1.26) | 0.74 (0.35, 1.58) |
| Apocrine metaplasia | **1.61 (1.01, 2.56)** | 0.94 (0.52, 1.71) | **4.35 (2.98, 6.36)*** | **2.10 (1.45, 3.06)*** | 1.03 (0.69, 1.54) | 0.74 (0.33, 1.66) | 1.74 (0.77, 3.93) | **2.35 (1.06, 5.20)*** | 0.77 (0.50, 1.18) | **2.61 (1.57, 4.34)*** | **4.87 (2.85, 8.33)*** | **4.86 (2.90, 8.16)*** |
| CCC | **0.55 (0.31, 0.96)** | **0.50 (0.25, 1.00)** | **0.32 (0.13, 0.74)*** | **0.31 (0.15, 0.63)*** | 0.72 (0.41, 1.27) | 1.14 (0.49, 2.68) | **0.44 (0.23, 0.85)** | **0.25 (0.12, 0.51)*** | 0.63 (0.35, 1.13) | 0.82 (0.43, 1.57) | **0.13 (0.02, 0.95)** | **0.22 (0.05, 0.92)** |
| UDH | 1.29 (0.69, 2.42) | 0.90 (0.42, 1.96) | 0.89 (0.48, 1.63) | **0.49 (0.27, 0.91)** | 0.90 (0.51, 1.59) | 1.10 (0.44, 2.76) | 0.48 (0.22, 1.02) | 0.59 (0.29, 1.21) | 0.94 (0.53, 1.69) | 0.96 (0.49, 1.86) | 1.41 (0.67, 2.98) | 0.54 (0.19, 1.54) |
| Sclerosing adenosis | 0.75 (0.51, 1.10) | **0.47 (0.28, 0.79)*** | 0.93 (0.61, 1.41) | 0.79 (0.54, 1.15) | 1.14 (0.78, 1.66) | 0.66 (0.29, 1.48) | 0.61 (0.37, 1.01) | **0.50 (0.30, 0.84)*** | **0.59 (0.39, 0.88)** | 0.79 (0.48, 1.28) | 0.94 (0.51, 1.71) | 0.62 (0.32, 1.21) |
| Fibrocystic change | 0.82 (0.54, 1.25) | 0.68 (0.41, 1.14) | 0.98 (0.64, 1.49) | 0.97 (0.66, 1.42) | 0.69 (0.46, 1.04) | 0.85 (0.44, 1.68) | 0.61 (0.36, 1.05) | **0.53 (0.31, 0.91)*** | 0.74 (0.49, 1.12) | 0.93 (0.57, 1.52) | 1.10 (0.60, 2.02) | 0.96 (0.53, 1.74) |
| Any (binary) | **0.71 (0.59, 0.85)*** | **0.65 (0.52, 0.81)*** | 0.89 (0.74, 1.08) | **0.83 (0.71, 0.98)** | **0.84 (0.70, 0.99)** | **0.67 (0.49, 0.93)** | 0.92 (0.72, 1.19) | **0.64 (0.49, 0.82)*** | 0.97 (0.81, 1.15) | 1.16 (0.94, 1.42) | **1.33 (1.02, 1.73)** | **0.66 (0.50, 0.87)*** |
| Any (continuous) | **0.8 (0.80, 0.98)*** | **0.77 (0.68, 0.88)*** | 0.95 (0.86, 1.06) | **0.89 (0.81, 0.99)** | 0.93 (0.84, 1.03) | **0.81 (0.66, 0.98)** | 0.89 (0.78, 1.02) | **0.75 (0.65, 0.86)*** | 0.92 (0.83, 1.02) | 1.06 (0.95, 1.19) | 1.09 (0.95, 1.26) | **0.81 (0.68, 0.96)*** |
| Early neoplastic, papillary and complex sclerosing lesions | | | | | | | | | | | | |
| DCIS | **0.67 (0.56, 0.80)*** | **0.52 (0.42, 0.65)*** | **0.72 (0.59, 0.87)*** | **0.80 (0.67, 0.94)*** | **0.73 (0.61, 0.86)*** | **0.53 (0.38, 0.73)*** | 0.83 (0.65, 1.08) | **0.60 (0.46, 0.78)*** | 1.00 (0.84, 1.19) | 1.16 (0.94, 1.42) | 1.07 (0.82, 1.40) | **0.43 (0.31, 0.59)*** |
| ADH | 0.6 (0.25, 1.52) | 0.25 (0.05, 1.16) | 0.52 (0.15, 1.77) | 0.61 (0.22, 1.67) | 0.77 (0.29, 2.01) | 1.64 (0.44, 6.17) | **0.35 (0.12, 0.97)** | **0.18 (0.06, 0.57)*** | 0.36 (0.12, 1.11) | 0.40 (0.09, 1.77) | 0.39 (0.05, 2.96) | 0.69 (0.16, 3.03) |
| FEA | **0.24 (0.06, 0.96)** | - | 0.87 (0.18, 4.24) | 0.26 (0.03, 2.07) | **0.12 (0.01, 0.95)** | - | 0.49 (0.11, 2.07) | **0.08 (0.01, 0.73)*** | - | 1.50 (0.37, 6.08) | - | - |
| Radial scar/complex sclerosing | 0.66 (0.24, 1.80) | 0.44 (0.11, 1.76) | - | 0.39 (0.11, 1.35) | 1.27 (0.49, 3.33) | 0.60 (0.07, 5.02) | 1.00 (0.28, 3.64) | 0.33 (0.07, 1.48) | 0.98 (0.36, 2.62) | 0.18 (0.02, 1.33) | - | - |
| Intraductal papilloma | **0.51 (0.31, 0.86)*** | **0.28 (0.13, 0.61)*** | 1.29 (0.75, 2.21) | 1.30 (0.79, 2.15) | 0.59 (0.34, 1.02) | 0.43 (0.13, 1.43) | **0.36 (0.19, 0.69)*** | **0.35 (0.18, 0.66)*** | **0.52 (0.29, 0.93)** | 0.45 (0.19, 1.07) | 0.89 (0.37, 2.13) | 1.48 (0.75, 2.93) |
| Any (binary) | **0.64 (0.53, 0.76)*** | **0.49 (0.40, 0.61)*** | **0.73 (0.61, 0.88)*** | **0.81 (0.69, 0.96)*** | **0.73 (0.62, 0.86)*** | **0.51 (0.37, 0.70)*** | 0.80 (0.62, 1.04) | **0.57 (0.45, 0.74)*** | 0.95 (0.80, 1.13) | 1.08 (0.88, 1.33) | 1.04 (0.80, 1.36) | **0.45 (0.34, 0.61)*** |
| Any (continuous) | **0.66 (0.56, 0.78)*** | **0.50 (0.41, 0.61)*** | **0.75 (0.63, 0.89)*** | **0.82 (0.71, 0.95)*** | **0.73 (0.63, 0.85)*** | **0.56 (0.41, 0.75)*** | **0.75 (0.60, 0.93)** | **0.54 (0.43, 0.67)*** | 0.92 (0.78, 1.07) | 1.03 (0.86, 1.23) | 0.98 (0.77, 1.24) | **0.50 (0.38, 0.66)*** |

**Supplementary Table 3.** Association between breast features and tumor characteristics in 1,756 breast cancer cases with reports from excision procedures, diagnosed before 2010. Odds ratios (ORs) and 95% confidence intervals (CIs) are presented from multinomial logistic regression models evaluating the association between breast features and tumor characteristics at diagnosis. Each OR reflects the odds of having a specific tumor characteristic versus a reference category, given the presence of a feature. All models are adjusted for potential confounders, including age at diagnosis, year of diagnosis, ethnicity, family history of breast cancer, menopausal status, and parity. All tests were two-sided. Statistically significant associations ($p < 0.05$) are indicated in bold. Multiple comparisons were controlled using the Benjamini–Hochberg procedure (false discovery rate adjustment); associations remaining significant after correction are denoted by *. LCIS: Lobular carcinoma in situ; ALH: Atypical lobular hyperplasia; CCC: Columnar cell change; UDH: Usual ductal hyperplasia; DCIS: Ductal carcinoma in situ; ADH: Atypical ductal hyperplasia; FEA: Flat epithelial atypia

| Reference | Stage I (n=621) | | ER-pos (n=1,098) | PR-pos (n=950) | Tumor size <2cm (n=754) | | Well-differentiated (n=250) | | Nodal status neg (n=881) | Luminal A (n=787) | | |
|---|---|---|---|---|---|---|---|---|---|---|---|---|
| | Stage II (n=798) | Stage III (n=337) | ER-neg (n=520) | PR-neg (n=657) | 2-5cm (n=638) | >5cm (n=124) | Moderately- (n=636) | Poorly- (n=764) | Positive (n=609) | Luminal B (n=223) | HER2-enriched (n=171) | Triple-neg (n=193) |
| Lobular neoplasia | | | | | | | | | | | | |
| LCIS | 0.69 (0.42, 1.12) | 0.73 (0.39, 1.37) | **0.40 (0.22, 0.73)*** | **0.50 (0.30, 0.82)*** | 0.94 (0.57, 1.55) | **2.18 (1.07, 4.45)** | 1.13 (0.63, 2.04) | **0.26 (0.12, 0.54)*** | 0.82 (0.50, 1.35) | **0.27 (0.11, 0.68)** | **0.20 (0.06, 0.66)*** | **0.30 (0.12, 0.77)*** |
| ALH | 0.46 (0.21, 1.01) | 0.34 (0.10, 1.17) | **0.23 (0.07, 0.76)** | 0.49 (0.22, 1.12) | **0.40 (0.16, 0.96)** | 0.67 (0.15, 3.00) | **0.31 (0.12, 0.77)*** | **0.20 (0.07, 0.53)*** | 0.88 (0.40, 1.94) | 1.23 (0.48, 3.18) | 0.27 (0.04, 2.07) | 0.20 (0.03, 1.55) |
| Any (binary) | **0.61 (0.39, 0.95)** | 0.56 (0.30, 1.03) | **0.33 (0.18, 0.58)*** | **0.49 (0.31, 0.78)*** | 0.77 (0.48, 1.22) | 1.61 (0.81, 3.21) | 0.78 (0.46, 1.30) | **0.20 (0.11, 0.39)*** | 0.76 (0.48, 1.21) | **0.37 (0.17, 0.78)** | **0.18 (0.05, 0.57)*** | **0.26 (0.10, 0.65)*** |
| Any (continuous) | **0.67 (0.46, 0.98)** | 0.66 (0.40, 1.10) | **0.42 (0.25, 0.69)*** | **0.56 (0.38, 0.83)*** | 0.78 (0.53, 1.16) | 1.47 (0.84, 2.58) | 0.83 (0.54, 1.27) | **0.29 (0.17, 0.51)*** | 0.86 (0.59, 1.27) | **0.54 (0.30, 0.99)** | **0.28 (0.10, 0.73)*** | **0.35 (0.16, 0.77)*** |
| Benign or non-atypical proliferative breast changes | | | | | | | | | | | | |
| Fibroadenoma | 0.73 (0.50, 1.06) | 0.79 (0.49, 1.27) | 0.71 (0.48, 1.05) | 0.78 (0.54, 1.12) | 0.91 (0.63, 1.30) | **0.21 (0.07, 0.69)*** | **0.57 (0.35, 0.92)** | **0.62 (0.39, 0.99)** | **0.61 (0.41, 0.90)*** | 0.83 (0.50, 1.40) | 0.52 (0.27, 1.03) | 0.85 (0.50, 1.46) |
| Calcification | **0.56 (0.43, 0.73)*** | **0.32 (0.21, 0.48)*** | 0.88 (0.67, 1.16) | 0.92 (0.71, 1.19) | **0.55 (0.41, 0.72)*** | **0.49 (0.28, 0.85)*** | **0.61 (0.42, 0.87)*** | **0.61 (0.43, 0.86)*** | **0.67 (0.50, 0.88)*** | 0.87 (0.59, 1.28) | 1.09 (0.72, 1.63) | 0.70 (0.45, 1.08) |
| Cyst | 0.80 (0.57, 1.11) | 0.63 (0.40, 1.00) | 1.02 (0.73, 1.43) | 0.91 (0.66, 1.26) | 0.88 (0.63, 1.23) | 0.71 (0.37, 1.36) | 0.70 (0.45, 1.09) | 0.73 (0.47, 1.12) | **0.63 (0.45, 0.90)*** | 0.66 (0.40, 1.11) | 0.82 (0.48, 1.40) | 0.96 (0.58, 1.56) |
| Apocrine metaplasia | **0.64 (0.43, 0.95)*** | **0.49 (0.28, 0.86)*** | 1.18 (0.80, 1.74) | 0.79 (0.53, 1.16) | **0.64 (0.42, 0.96)** | 0.72 (0.34, 1.52) | 0.73 (0.44, 1.22) | 0.65 (0.39, 1.08) | 0.91 (0.61, 1.36) | 0.80 (0.44, 1.43) | 0.88 (0.47, 1.65) | 0.93 (0.52, 1.69) |
| CCC | **0.53 (0.33, 0.84)*** | **0.50 (0.26, 0.95)** | 0.80 (0.50, 1.30) | 0.78 (0.50, 1.23) | 0.81 (0.51, 1.27) | 0.47 (0.16, 1.34) | **0.48 (0.27, 0.85)*** | **0.42 (0.24, 0.74)*** | **0.56 (0.34, 0.92)** | 0.78 (0.41, 1.50) | 0.70 (0.33, 1.52) | 0.90 (0.46, 1.77) |
| UDH | **0.62 (0.42, 0.92)*** | **0.16 (0.07, 0.37)*** | 0.96 (0.63, 1.45) | 0.71 (0.47, 1.07) | **0.60 (0.39, 0.91)** | **0.30 (0.11, 0.86)** | **0.53 (0.31, 0.88)*** | **0.51 (0.31, 0.85)*** | **0.52 (0.33, 0.81)*** | 0.78 (0.43, 1.40) | 0.94 (0.50, 1.76) | 0.67 (0.34, 1.30) |
| Sclerosing adenosis | **0.53 (0.36, 0.79)*** | **0.39 (0.21, 0.70)*** | 1.25 (0.85, 1.85) | 0.98 (0.66, 1.44) | **0.51 (0.33, 0.78)*** | **0.40 (0.17, 0.97)** | **0.46 (0.28, 0.77)*** | **0.49 (0.30, 0.80)*** | 0.71 (0.46, 1.08) | 0.83 (0.46, 1.50) | 1.42 (0.81, 2.49) | 1.08 (0.59, 1.96) |
| Fibrocystic change | 0.79 (0.62, 1.00) | **0.49 (0.34, 0.69)*** | 0.86 (0.67, 1.11) | 0.82 (0.64, 1.04) | 0.81 (0.63, 1.04) | **0.47 (0.28, 0.81)*** | **0.65 (0.46, 0.90)*** | **0.60 (0.44, 0.83)*** | **0.65 (0.50, 0.84)*** | 0.87 (0.61, 1.24) | 0.78 (0.52, 1.18) | 0.81 (0.55, 1.18) |
| Any (binary) | **0.70 (0.56, 0.87)*** | **0.46 (0.35, 0.61)*** | 0.92 (0.74, 1.14) | 0.85 (0.70, 1.05) | **0.74 (0.59, 0.91)*** | **0.45 (0.30, 0.68)*** | **0.71 (0.53, 0.96)** | **0.68 (0.51, 0.92)*** | **0.67 (0.54, 0.83)*** | 0.96 (0.71, 1.30) | 0.92 (0.66, 1.29) | 0.88 (0.63, 1.21) |
| Any (continuous) | **0.86 (0.80, 0.92)*** | **0.73 (0.65, 0.82)*** | 0.97 (0.90, 1.05) | 0.94 (0.87, 1.01) | **0.88 (0.81, 0.95)*** | **0.74 (0.62, 0.88)*** | **0.83 (0.76, 0.91)*** | **0.82 (0.75, 0.90)*** | **0.85 (0.78, 0.92)*** | 0.93 (0.83, 1.03) | 0.95 (0.85, 1.07) | 0.93 (0.83, 1.05) |
| Early neoplastic, papillary and complex sclerosing lesions | | | | | | | | | | | | |
| DCIS | **0.53 (0.42, 0.66)*** | **0.58 (0.43, 0.77)*** | 0.89 (0.72, 1.11) | **0.75 (0.61, 0.93)*** | **0.64 (0.51, 0.80)*** | **0.52 (0.35, 0.77)*** | 1.10 (0.81, 1.50) | 0.89 (0.66, 1.20) | 1.13 (0.90, 1.41) | 1.11 (0.80, 1.53) | **1.97 (1.32, 2.95)*** | **0.51 (0.37, 0.71)*** |
| ADH | **0.53 (0.31, 0.93)*** | **0.21 (0.07, 0.61)*** | **0.39 (0.19, 0.81)** | 0.68 (0.38, 1.20) | **0.53 (0.30, 0.94)** | 0.15 (0.02, 1.09) | 0.56 (0.30, 1.02) | **0.19 (0.09, 0.40)*** | **0.52 (0.28, 0.97)** | 0.82 (0.39, 1.74) | 0.50 (0.18, 1.43) | 0.32 (0.10, 1.05) |
| FEA | 1.15 (0.25, 5.32) | 0.68 (0.07, 6.84) | 1.30 (0.30, 5.57) | 0.90 (0.21, 3.88) | 0.75 (0.17, 3.25) | - | 0.45 (0.06, 3.27) | 0.54 (0.09, 3.32) | 0.53 (0.10, 2.74) | 0.87 (0.10, 7.88) | - | 3.26 (0.69, 15.35) |
| Radial scar/complex sclerosing | 0.61 (0.31, 1.19) | 0.51 (0.20, 1.31) | 0.57 (0.27, 1.21) | 0.51 (0.25, 1.03) | 0.98 (0.51, 1.87) | 0.84 (0.24, 2.94) | **0.37 (0.17, 0.81)*** | **0.39 (0.18, 0.84)*** | **0.45 (0.21, 0.94)** | 0.70 (0.26, 1.87) | 0.54 (0.16, 1.81) | 0.68 (0.23, 2.00) |
| Intraductal papilloma | 0.68 (0.37, 1.24) | 0.75 (0.36, 1.60) | 0.68 (0.36, 1.29) | 0.60 (0.33, 1.10) | 0.88 (0.50, 1.58) | 1.00 (0.37, 2.76) | 0.97 (0.44, 2.11) | 0.78 (0.36, 1.70) | 0.95 (0.52, 1.73) | 0.89 (0.40, 1.97) | 0.86 (0.35, 2.11) | 0.34 (0.10, 1.14) |
| Any (binary) | **0.51 (0.41, 0.64)*** | **0.57 (0.43, 0.76)*** | 0.89 (0.71, 1.11) | **0.74 (0.60, 0.91)*** | **0.63 (0.50, 0.79)*** | **0.53 (0.35, 0.79)*** | 1.08 (0.79, 1.47) | 0.84 (0.62, 1.13) | 1.11 (0.89, 1.39) | 1.02 (0.74, 1.42) | **1.81 (1.20, 2.72)*** | **0.50 (0.36, 0.70)*** |
| Any (continuous) | **0.60 (0.50, 0.71)*** | **0.60 (0.48, 0.75)*** | **0.82 (0.69, 0.97)** | **0.75 (0.63, 0.88)*** | **0.71 (0.59, 0.85)*** | **0.57 (0.41, 0.79)*** | 0.89 (0.70, 1.12) | **0.71 (0.56, 0.90)*** | 0.95 (0.80, 1.13) | 1.00 (0.79, 1.27) | 1.23 (0.95, 1.60) | **0.54 (0.40, 0.71)*** |

**Supplementary Table 4.** Association between breast features and tumor characteristics in 2,232 breast cancer cases with reports from excision procedures, diagnosed 2010 and after. Odds ratios (ORs) and 95% confidence intervals (CIs) are presented from multinomial logistic regression models evaluating the association between breast features and tumor characteristics at diagnosis. Each OR reflects the odds of having a specific tumor characteristic versus a reference category, given the presence of a feature. All models are adjusted for potential confounders, including age at diagnosis, year of diagnosis, ethnicity, family history of breast cancer, menopausal status, and parity. All tests were two-sided. Statistically significant associations ($p < 0.05$) are indicated in bold. Multiple comparisons were controlled using the Benjamini–Hochberg procedure (false discovery rate adjustment); associations remaining significant after correction are denoted by *. LCIS: Lobular carcinoma in situ; ALH: Atypical lobular hyperplasia; CCC: Columnar cell change; UDH: Usual ductal hyperplasia; DCIS: Ductal carcinoma in situ; ADH: Atypical ductal hyperplasia; FEA: Flat epithelial atypia

| Reference | Stage I (n=983) | | ER-pos (n=1,773) | PR-pos (n=1,479) | Tumor size <2cm (n=1,090) | | Well-differentiated (n=317) | | Nodal status neg (n=1,373) | Luminal A (n=1,436) | | |
|---|---|---|---|---|---|---|---|---|---|---|---|---|
| | Stage II (n=836) | Stage III (n=413) | ER-neg (n=444) | PR-neg (n=733) | 2-5cm (n=872) | >5cm (n=160) | Moderately- (n=927) | Poorly- (n=910) | Positive (n=662) | Luminal B (n=334) | HER2-enriched (n=199) | Triple-neg (n=197) |
| Lobular neoplasia | | | | | | | | | | | | |
| LCIS | 0.92 (0.64, 1.32) | 0.82 (0.51, 1.31) | **0.46 (0.27, 0.77)*** | 0.71 (0.49, 1.04) | 0.89 (0.63, 1.28) | 1.50 (0.86, 2.64) | 1.50 (0.93, 2.43) | **0.50 (0.29, 0.86)*** | 0.90 (0.62, 1.31) | **0.47 (0.27, 0.81)*** | 0.50 (0.25, 1.01) | **0.45 (0.21, 0.94)** |
| ALH | **0.29 (0.14, 0.61)*** | **0.27 (0.10, 0.70)*** | **0.17 (0.04, 0.70)*** | 0.62 (0.31, 1.23) | **0.35 (0.18, 0.71)*** | 0.31 (0.07, 1.33) | 0.80 (0.38, 1.69) | **0.34 (0.14, 0.82)*** | **0.45 (0.21, 0.96)** | **0.28 (0.09, 0.92)** | 0.17 (0.02, 1.23) | 0.17 (0.02, 1.24) |
| Any (binary) | 0.85 (0.60, 1.19) | 0.76 (0.49, 1.18) | **0.41 (0.25, 0.68)*** | **0.66 (0.46, 0.95)** | 0.86 (0.62, 1.20) | 1.29 (0.75, 2.22) | 1.16 (0.76, 1.79) | **0.45 (0.28, 0.73)*** | 0.87 (0.61, 1.24) | **0.45 (0.27, 0.76)*** | **0.42 (0.21, 0.85)** | **0.43 (0.21, 0.86)** |
| Any (continuous) | **0.75 (0.56, 1.00)** | **0.67 (0.46, 0.99)** | **0.44 (0.28, 0.70)*** | **0.73 (0.54, 0.99)** | 0.76 (0.57, 1.01) | 1.06 (0.67, 1.70) | 1.22 (0.84, 1.76) | **0.50 (0.32, 0.77)*** | 0.80 (0.59, 1.09) | **0.48 (0.30, 0.76)*** | **0.47 (0.26, 0.88)** | **0.43 (0.23, 0.83)*** |
| Benign or non-atypical proliferative breast changes | | | | | | | | | | | | |
| Fibroadenoma | 1.21 (0.92, 1.58) | **1.43 (1.04, 1.98)*** | 0.96 (0.72, 1.30) | 0.90 (0.69, 1.16) | **1.32 (1.02, 1.70)** | 1.47 (0.94, 2.28) | 1.09 (0.75, 1.58) | 1.18 (0.81, 1.71) | 1.14 (0.88, 1.49) | 0.74 (0.52, 1.05) | 1.03 (0.69, 1.55) | 0.72 (0.45, 1.14) |
| Calcification | **0.64 (0.52, 0.79)*** | **0.77 (0.59, 1.00)** | 0.94 (0.75, 1.19) | 1.03 (0.85, 1.26) | **0.71 (0.58, 0.87)*** | 0.76 (0.52, 1.11) | 1.27 (0.96, 1.69) | 1.02 (0.76, 1.36) | **0.68 (0.54, 0.84)*** | 1.16 (0.90, 1.50) | 1.28 (0.94, 1.76) | **0.65 (0.45, 0.93)** |
| Cyst | 1.11 (0.87, 1.42) | 1.00 (0.74, 1.35) | 0.95 (0.73, 1.25) | 0.82 (0.64, 1.04) | 0.95 (0.75, 1.20) | 1.44 (0.97, 2.14) | 0.89 (0.64, 1.23) | 0.81 (0.58, 1.12) | 1.05 (0.83, 1.34) | 0.92 (0.68, 1.26) | 0.94 (0.64, 1.38) | 0.93 (0.63, 1.38) |
| Apocrine metaplasia | 0.76 (0.58, 1.00) | **0.46 (0.30, 0.69)*** | **1.70 (1.28, 2.26)*** | **1.36 (1.05, 1.77)** | **0.68 (0.52, 0.90)*** | 0.72 (0.43, 1.23) | 1.15 (0.77, 1.74) | 1.33 (0.89, 2.00) | **0.70 (0.52, 0.95)** | 1.03 (0.71, 1.49) | 1.47 (0.97, 2.23) | **1.72 (1.15, 2.56)*** |
| CCC | **0.71 (0.53, 0.95)*** | **0.50 (0.33, 0.76)*** | 0.95 (0.67, 1.33) | 0.89 (0.67, 1.20) | **0.69 (0.52, 0.93)*** | **0.55 (0.30, 1.00)** | **0.67 (0.47, 0.97)** | **0.51 (0.35, 0.75)*** | **0.65 (0.47, 0.90)*** | 1.02 (0.70, 1.48) | 0.99 (0.61, 1.60) | 0.87 (0.53, 1.45) |
| UDH | 0.82 (0.62, 1.07) | **0.50 (0.34, 0.75)*** | 0.81 (0.58, 1.13) | 0.95 (0.72, 1.24) | 0.85 (0.65, 1.11) | **0.43 (0.23, 0.81)*** | 0.89 (0.61, 1.29) | 0.90 (0.62, 1.32) | **0.62 (0.46, 0.84)*** | 1.16 (0.82, 1.63) | 0.87 (0.55, 1.38) | 0.76 (0.47, 1.24) |
| Sclerosing adenosis | 0.80 (0.59, 1.07) | **0.59 (0.39, 0.89)*** | 0.85 (0.60, 1.20) | 0.92 (0.69, 1.23) | 0.76 (0.56, 1.02) | 0.73 (0.41, 1.29) | 1.13 (0.74, 1.72) | 1.11 (0.72, 1.69) | 0.73 (0.52, 1.01) | 1.28 (0.89, 1.82) | 1.07 (0.67, 1.71) | 0.66 (0.38, 1.16) |
| Fibrocystic change | **0.72 (0.59, 0.89)*** | **0.69 (0.53, 0.91)*** | 0.95 (0.75, 1.20) | 0.88 (0.72, 1.08) | **0.73 (0.60, 0.90)*** | 0.80 (0.54, 1.18) | 0.87 (0.65, 1.15) | 0.87 (0.66, 1.16) | **0.76 (0.61, 0.95)** | 0.83 (0.63, 1.09) | 1.01 (0.73, 1.41) | 0.76 (0.53, 1.08) |
| Any (binary) | **0.82 (0.68, 0.99)** | 0.84 (0.66, 1.08) | 1.13 (0.92, 1.41) | 1.12 (0.93, 1.34) | **0.78 (0.65, 0.94)*** | 0.94 (0.66, 1.33) | 0.94 (0.72, 1.22) | 0.93 (0.72, 1.22) | **0.79 (0.65, 0.96)** | 1.02 (0.80, 1.31) | **1.46 (1.06, 2.00)** | 0.91 (0.67, 1.23) |
| Any (continuous) | **0.92 (0.87, 0.97)*** | **0.88 (0.82, 0.95)*** | 1.00 (0.94, 1.06) | 0.98 (0.93, 1.04) | **0.92 (0.87, 0.97)*** | 0.93 (0.84, 1.03) | 0.99 (0.92, 1.07) | 0.97 (0.90, 1.05) | **0.91 (0.85, 0.96)*** | 1.00 (0.93, 1.07) | 1.03 (0.95, 1.13) | 0.93 (0.84, 1.02) |
| Early neoplastic, papillary and complex sclerosing lesions | | | | | | | | | | | | |
| DCIS | **0.71 (0.59, 0.87)*** | **0.74 (0.57, 0.94)*** | 0.92 (0.74, 1.15) | 0.90 (0.74, 1.08) | **0.81 (0.67, 0.98)** | **0.51 (0.36, 0.72)*** | 1.19 (0.91, 1.55) | 1.09 (0.83, 1.42) | 0.98 (0.80, 1.20) | **1.43 (1.10, 1.85)** | **1.58 (1.14, 2.19)** | **0.65 (0.48, 0.88)*** |
| ADH | 0.69 (0.45, 1.05) | **0.33 (0.17, 0.66)*** | **0.31 (0.15, 0.64)*** | **0.48 (0.29, 0.79)*** | 0.81 (0.54, 1.23) | **0.27 (0.08, 0.89)** | 0.66 (0.40, 1.09) | **0.33 (0.19, 0.58)*** | **0.56 (0.35, 0.91)** | 0.90 (0.53, 1.54) | **0.35 (0.13, 0.97)** | **0.27 (0.08, 0.86)** |
| FEA | 0.88 (0.55, 1.39) | **0.45 (0.24, 0.85)*** | **0.33 (0.16, 0.70)*** | - | 0.79 (0.50, 1.23) | 0.41 (0.16, 1.09) | 0.72 (0.42, 1.25) | **0.30 (0.16, 0.56)*** | 0.69 (0.43, 1.11) | 0.85 (0.48, 1.52) | 0.58 (0.24, 1.39) | **0.19 (0.05, 0.80)** |
| Radial scar/complex sclerosing | **0.60 (0.38, 0.97)** | 0.98 (0.58, 1.66) | 0.78 (0.46, 1.33) | 0.87 (0.56, 1.35) | 0.76 (0.50, 1.18) | 0.88 (0.40, 1.92) | 0.79 (0.45, 1.37) | 0.64 (0.36, 1.14) | 0.90 (0.56, 1.44) | 1.08 (0.62, 1.87) | 0.85 (0.40, 1.80) | 0.42 (0.15, 1.17) |
| Intraductal papilloma | **0.51 (0.36, 0.73)*** | **0.53 (0.34, 0.83)*** | **0.63 (0.40, 0.97)** | **0.64 (0.45, 0.91)** | **0.55 (0.39, 0.79)*** | 0.65 (0.35, 1.21) | 0.78 (0.51, 1.19) | **0.58 (0.37, 0.90)*** | 0.71 (0.49, 1.01) | 1.05 (0.68, 1.61) | **0.33 (0.14, 0.77)** | 0.94 (0.53, 1.65) |
| Any (binary) | **0.67 (0.55, 0.82)*** | **0.67 (0.52, 0.87)*** | **0.94 (0.75, 1.17)** | 0.88 (0.73, 1.07) | **0.79 (0.65, 0.96)*** | **0.49 (0.34, 0.69)*** | 1.04 (0.79, 1.38) | 0.88 (0.67, 1.16) | **0.87 (0.77, 0.99)** | **1.34 (1.02, 1.75)** | **1.52 (1.08, 2.14)** | **0.67 (0.49, 0.91)*** |
| Any (continuous) | **0.73 (0.65, 0.83)*** | **0.71 (0.60, 0.83)*** | **0.78 (0.67, 0.91)*** | **0.80 (0.70, 0.91)*** | **0.80 (0.71, 0.91)*** | **0.60 (0.47, 0.77)*** | 0.94 (0.80, 1.11) | **0.77 (0.65, 0.91)*** | 0.98 (0.80, 1.20) | 1.13 (0.97, 1.32) | 0.97 (0.79, 1.19) | **0.63 (0.50, 0.80)*** |

**Supplementary Table 5.** Association between coexisting features and 10-year overall survival in 3,164 breast cancer patients with CNB reports (613 events). Hazard ratios (HRs) with 95% confidence intervals (CIs) are presented from Cox proportional hazards models evaluating the impact of coexisting breast features on 10-year overall survival among breast cancer patients. All tests were two-sided. Statistically significant associations ($p < 0.05$) are indicated in bold. Multiple comparisons were controlled using the Benjamini–Hochberg procedure (false discovery rate adjustment); associations remaining significant after correction are denoted by *. a) Adjusted for age at diagnosis and year of diagnosis. b) Model one further adjusted for menstruation status, ethnicity, family history of cancer and parity. c) Model two further adjusted for tumour characteristics: stage and subtype.

| | | | Model 1[a] | Model 2[b] | Model 3[c] |
|---|---|---|---|---|---|
| | Number of patients | Number of events | HR (95% CI) | HR (95% CI) | HR (95% CI) |
| Lobular neoplasia | | | | | |
| Lobular carcinoma in situ (LCIS) | 60 (1.9%) | 10 (16.7%) | 0.96 (0.51, 1.79) | 0.98 (0.53, 1.84) | 1.07 (0.57, 2.01) |
| Atypical lobular hyperplasia (ALH) | 11 (0.3%) | 1 (9.1%) | 0.52 (0.07, 3.68) | 0.61 (0.09, 4.35) | 1.11 (0.15, 7.93) |
| Any (binary) | 66 (2.1%) | 11 (2.0%) | 0.97 (0.53, 1.76) | 1.00 (0.55, 1.83) | 1.12 (0.61, 2.03) |
| Any (continuous) | - | - | 0.90 (0.52, 1.57) | 0.94 (0.54, 1.65) | 1.07 (0.60, 1.91) |
| Benign or non-atypical proliferative breast changes | | | | | |
| Fibroadenoma | 109 (3.4%) | 15 (13.8%) | 0.79 (0.48, 1.33) | 0.83 (0.50, 1.39) | 0.96 (0.57, 1.61) |
| Calcification | 630 (19.9%) | 104 (16.5%) | 0.88 (0.71, 1.09) | 0.88 (0.71, 1.09) | 0.95 (0.77, 1.18) |
| Cyst | 84 (2.7%) | 9 (10.7%) | 0.55 (0.28, 1.06) | 0.53 (0.27, 1.02) | 0.70 (0.36, 1.35) |
| Apocrine metaplasia | 117 (3.7%) | 10 (8.5%) | **0.46 (0.25, 0.87)** | **0.49 (0.26, 0.92)** | **0.43 (0.23, 0.80)** |
| Columnar cell change | 65 (2.1%) | 5 (7.7%) | **0.41 (0.17, 1.00)** | 0.44 (0.18, 1.07) | 0.53 (0.22, 1.27) |
| Usual ductal hyperplasia (UDH) | 62 (2.0%) | 6 (9.7%) | 0.66 (0.29, 1.47) | 0.67 (0.30, 1.50) | 0.68 (0.30, 1.52) |
| Sclerosing adenosis | 136 (4.3%) | 18 (13.2%) | 0.71 (0.44, 1.14) | 0.74 (0.46, 1.19) | 0.87 (0.54, 1.39) |
| Fibrocystic change | 125 (4.0%) | 22 (17.6%) | 0.98 (0.64, 1.50) | 0.98 (0.64, 1.50) | 1.03 (0.67, 1.59) |
| Any (binary) | 942 (30.0%) | 141 (15.0%) | **0.75 (0.62, 0.90)*** | **0.75 (0.62, 0.91)*** | **0.82 (0.68, 1.00)** |
| Any (continuous) | - | - | **0.84 (0.74, 0.95)*** | **0.85 (0.75, 0.96)** | 0.90 (0.79, 1.02) |
| Early neoplastic, papillary and complex sclerosing lesions | | | | | |
| Ductal carcinoma in situ (DCIS) | 942 (29.8%) | 145 (15.4%) | **0.79 (0.65, 0.95)*** | **0.81 (0.67, 0.98)** | 0.94 (0.78, 1.14) |
| Atypical ductal hyperplasia (ADH) | 21 (0.7%) | 2 (9.5%) | 0.41 (0.10, 1.63) | 0.45 (0.11, 1.81) | 0.53 (0.13, 2.12) |
| Flat epithelial atypia (FEA) | 9 (0.3%) | 0 (0.0%) | - | - | - |
| Radial scar or complex sclerosing lesion | 19 (0.6%) | 1 (5.3%) | 0.33 (0.05, 2.37) | 0.33 (0.05, 2.35) | 0.36 (0.05, 2.58) |
| Intraductal papilloma | 70 (2.2%) | 5 (7.1%) | **0.36 (0.15, 0.86)** | **0.37 (0.15, 0.90)** | 0.44 (0.18, 1.05) |
| Any (binary) | 993 (31.4%) | 149 (15.0%) | **0.75 (0.63, 0.91)*** | **0.78 (0.65, 0.94)** | 0.91 (0.75, 1.10) |
| Any (continuous) | - | - | **0.74 (0.63, 0.88)*** | **0.77 (0.64, 0.91)*** | 0.88 (0.74, 1.05) |

**Supplementary Table 6.** Association between coexisting features and 10-year overall survival in 1,756 breast cancer cases with reports from excision procedures, diagnosed before 2010. Hazard ratios (HRs) with 95% confidence intervals (CIs) are presented from Cox proportional hazards models evaluating the impact of coexisting breast features on 10-year overall survival among breast cancer patients. All tests were two-sided. Statistically significant associations ($p < 0.05$) are indicated in bold. Multiple comparisons were controlled using the Benjamini–Hochberg procedure (false discovery rate adjustment); associations remaining significant after correction are denoted by *. a) Adjusted for age at diagnosis and year of diagnosis. b) Model one further adjusted for menstruation status, ethnicity, family history of cancer and parity. c) Model two further adjusted for tumour characteristics: stage and subtype.

| | Number of patients | Number of events | Model 1[a] HR (95% CI) | Model 2[b] HR (95% CI) | Model 3[c] HR (95% CI) |
|---|---|---|---|---|---|
| Lobular neoplasia | | | | | |
| Lobular carcinoma in situ (LCIS) | 86 (4.9%) | 18 (20.9%) | 1.05 (0.65, 1.69) | 1.01 (0.63, 1.63) | 1.08 (0.67, 1.75) |
| Atypical lobular hyperplasia (ALH) | 31 (1.8%) | 4 (12.9%) | 0.69 (0.26, 1.84) | 0.71 (0.26, 1.90) | 0.83 (0.30, 2.25) |
| Any (binary) | 102 (5.8%) | 19 (18.6%) | 0.95 (0.60, 1.51) | 0.92 (0.58, 1.47) | 1.04 (0.65, 1.67) |
| Any (continuous) | - | - | 0.96 (0.66, 1.41) | 0.95 (0.64, 1.40) | 1.02 (0.69, 1.49) |
| Benign or non-atypical proliferative breast changes | | | | | |
| Fibroadenoma | 152 (8.7%) | 25 (16.4%) | 0.88 (0.58, 1.32) | 0.90 (0.60, 1.36) | 0.97 (0.64, 1.47) |
| Calcification | 321 (18.3%) | 47 (14.6%) | **0.72 (0.53, 0.98)** | **0.71 (0.52, 0.97)** | 0.89 (0.65, 1.22) |
| Cyst | 194 (11.0%) | 25 (12.9%) | 0.70 (0.46, 1.05) | 0.70 (0.46, 1.06) | 0.74 (0.49, 1.12) |
| Apocrine metaplasia | 135 (7.7%) | 16 (11.9%) | 0.62 (0.37, 1.02) | 0.63 (0.38, 1.04) | 0.71 (0.43, 1.18) |
| Columnar cell change | 95 (5.4%) | 8 (8.4%) | **0.47 (0.23, 0.95)** | **0.49 (0.24, 0.99)** | 0.50 (0.25, 1.03) |
| Usual ductal hyperplasia (UDH) | 122 (6.9%) | 6 (4.9%) | **0.24 (0.11, 0.55)*** | **0.25 (0.11, 0.56)*** | **0.34 (0.15, 0.77)** |
| Sclerosing adenosis | 128 (7.3%) | 16 (12.5%) | 0.70 (0.42, 1.16) | 0.72 (0.44, 1.20) | 0.92 (0.55, 1.53) |
| Fibrocystic change | 415 (23.6%) | 59 (14.2%) | **0.71 (0.54, 0.94)** | 0.76 (0.57, 1.01) | 0.87 (0.65, 1.16) |
| Any (binary) | 743 (42.3%) | 117 (15.7%) | **0.72 (0.57, 0.90)*** | **0.73 (0.58, 0.91)*** | 0.85 (0.67, 1.06) |
| Any (continuous) | - | - | **0.84 (0.76, 0.93)*** | **0.85 (0.77, 0.94)*** | **0.90 (0.82, 1.00)** |
| Early neoplastic, papillary and complex sclerosing lesions | | | | | |
| Ductal carcinoma in situ (DCIS) | 1074 (61.2%) | 191 (17.8%) | 0.84 (0.68, 1.05) | 0.86 (0.69, 1.07) | 1.02 (0.81, 1.28) |
| Atypical ductal hyperplasia (ADH) | 60 (3.4%) | 8 (13.3%) | 0.69 (0.34, 1.40) | 0.72 (0.35, 1.45) | 0.94 (0.46, 1.92) |
| Flat epithelial atypia (FEA) | 8 (0.5%) | 1 (12.5%) | 0.71 (0.10, 5.08) | 0.70 (0.10, 5.03) | 0.80 (0.11, 5.76) |
| Radial scar or complex sclerosing lesion | 43 (2.4%) | 9 (20.9%) | 1.08 (0.56, 2.09) | 1.06 (0.54, 2.05) | 1.10 (0.57, 2.14) |
| Intraductal papilloma | 57 (3.2%) | 11 (19.3%) | 1.04 (0.57, 1.89) | 1.05 (0.57, 1.93) | 1.12 (0.60, 2.07) |
| Any (binary) | 1108 (63.1%) | 197 (17.8%) | 0.84 (0.67, 1.04) | 0.85 (0.68, 1.06) | 0.99 (0.79, 1.25) |
| Any (continuous) | - | - | 0.88 (0.73, 1.05) | 0.89 (0.74, 1.06) | 1.02 (0.85, 1.23) |

**Supplementary Table 7.** Association between coexisting features and 10-year overall survival in 2,232 breast cancer cases with reports from excision procedures, diagnosed 2010 and after. Hazard ratios (HRs) with 95% confidence intervals (CIs) are presented from Cox proportional hazards models evaluating the impact of coexisting breast features on 10-year overall survival among breast cancer patients. All tests were two-sided. Statistically significant associations ($p < 0.05$) are indicated in bold. Multiple comparisons were controlled using the Benjamini–Hochberg procedure (false discovery rate adjustment); associations remaining significant after correction are denoted by *. a) Adjusted for age at diagnosis and year of diagnosis. b) Model one further adjusted for menstruation status, ethnicity, family history of cancer and parity. c) Model two further adjusted for tumour characteristics: stage and subtype.

| | Number of patients | Number of events | Model 1[a] HR (95% CI) | Model 2[b] HR (95% CI) | Model 3[c] HR (95% CI) |
|---|---|---|---|---|---|
| Lobular neoplasia | | | | | |
| Lobular carcinoma in situ (LCIS) | 160 (7.2%) | 18 (11.2%) | 1.01 (0.62, 1.63) | 1.01 (0.62, 1.63) | 1.17 (0.72, 1.90) |
| Atypical lobular hyperplasia (ALH) | 49 (2.2%) | 4 (8.2%) | 0.86 (0.32, 2.31) | 0.79 (0.29, 2.12) | 0.93 (0.34, 2.52) |
| Any (binary) | 185 (8.3%) | 22 (11.9%) | 1.11 (0.72, 1.71) | 1.10 (0.71, 1.71) | 1.23 (0.79, 1.92) |
| Any (continuous) | - | - | 0.98 (0.66, 1.45) | 0.96 (0.65, 1.43) | 1.10 (0.73, 1.65) |
| Benign or non-atypical proliferative breast changes | | | | | |
| Fibroadenoma | 339 (15.2%) | 42 (12.4%) | 0.98 (0.71, 1.37) | 0.92 (0.66, 1.27) | 0.94 (0.67, 1.31) |
| Calcification | 699 (31.3%) | 69 (9.9%) | **0.74 (0.56, 0.97)** | **0.71 (0.54, 0.93)** | **0.72 (0.55, 0.95)** |
| Cyst | 432 (19.4%) | 49 (11.3%) | 0.91 (0.67, 1.24) | 0.84 (0.62, 1.16) | 0.88 (0.64, 1.21) |
| Apocrine metaplasia | 288 (12.9%) | 32 (11.1%) | 0.89 (0.62, 1.29) | 0.89 (0.61, 1.29) | 1.00 (0.69, 1.46) |
| Columnar cell change | 253 (11.3%) | 28 (11.1%) | 0.93 (0.63, 1.38) | 0.92 (0.62, 1.36) | 1.06 (0.71, 1.58) |
| Usual ductal hyperplasia (UDH) | 291 (13.0%) | 39 (13.4%) | 1.09 (0.77, 1.52) | 1.03 (0.74, 1.45) | 1.17 (0.83, 1.66) |
| Sclerosing adenosis | 245 (11.0%) | 30 (12.2%) | 1.07 (0.73, 1.57) | 1.06 (0.73, 1.56) | 1.21 (0.82, 1.77) |
| Fibrocystic change | 627 (28.1%) | 69 (11.0%) | 0.91 (0.70, 1.20) | 0.91 (0.69, 1.19) | 0.99 (0.75, 1.30) |
| Any (binary) | 1324 (59.3%) | 158 (12.0%) | 0.90 (0.71, 1.13) | 0.86 (0.68, 1.09) | 0.90 (0.71, 1.14) |
| Any (continuous) | - | - | 0.96 (0.89, 1.04) | 0.95 (0.88, 1.02) | 0.98 (0.91, 1.06) |
| Early neoplastic, papillary and complex sclerosing lesions | | | | | |
| Ductal carcinoma in situ (DCIS) | 1375 (61.6%) | 176 (12.8%) | 1.00 (0.79, 1.27) | 0.98 (0.77, 1.25) | 1.10 (0.87, 1.41) |
| Atypical ductal hyperplasia (ADH) | 110 (4.9%) | 6 (5.5%) | **0.41 (0.18, 0.92)** | **0.41 (0.18, 0.91)** | 0.49 (0.22, 1.10) |
| Flat epithelial atypia (FEA) | 100 (4.5%) | 7 (7.0%) | 0.60 (0.28, 1.27) | 0.64 (0.30, 1.38) | 0.81 (0.37, 1.75) |
| Radial scar or complex sclerosing lesion | 105 (4.7%) | 8 (7.6%) | 0.60 (0.30, 1.22) | 0.61 (0.30, 1.23) | 0.59 (0.29, 1.21) |
| Intraductal papilloma | 183 (8.2%) | 30 (16.4%) | 1.22 (0.84, 1.78) | 1.22 (0.83, 1.78) | 1.46 (0.99, 2.15) |
| Any (binary) | 1480 (66.3%) | 196 (13.2%) | 1.10 (0.86, 1.41) | 1.10 (0.85, 1.41) | 1.24 (0.97, 1.60) |
| Any (continuous) | - | - | 0.93 (0.79, 1.08) | 0.92 (0.78, 1.08) | 1.02 (0.87, 1.20) |

**Supplementary Table 8.** Association between coexisting features and 10-year overall survival in 1,604 breast cancer cases with reports from excision procedures diagnosed with Stage I breast cancer. Hazard ratios (HRs) with 95% confidence intervals (CIs) are presented from Cox proportional hazards models evaluating the impact of coexisting breast features on 10-year overall survival among breast cancer patients. All tests were two-sided. Statistically significant associations ($p < 0.05$) are indicated in bold. Multiple comparisons were controlled using the Benjamini–Hochberg procedure (false discovery rate adjustment); associations remaining significant after correction are denoted by *. a) Adjusted for age at diagnosis and year of diagnosis. b) Model one further adjusted for menstruation status, ethnicity, family history of cancer and parity. c) Model two further adjusted for tumour characteristics: subtype.

| | Number of patients | Number of events | Model 1[a] HR (95% CI) | Model 2[b] HR (95% CI) | Model 3[c] HR (95% CI) |
|---|---|---|---|---|---|
| Lobular neoplasia | | | | | |
| Lobular carcinoma in situ (LCIS) | 109 (6.8%) | 6 (5.5%) | 0.81 (0.35, 1.83) | 0.79 (0.34, 1.80) | 0.78 (0.34, 1.80) |
| Atypical lobular hyperplasia (ALH) | 52 (3.2%) | 2 (3.8%) | 0.72 (0.18, 2.92) | 0.70 (0.17, 2.87) | 0.71 (0.17, 2.92) |
| Any (binary) | 134 (8.4%) | 8 (6.0%) | 0.92 (0.45, 1.88) | 0.91 (0.44, 1.87) | 0.91 (0.44, 1.88) |
| Any (continuous) | - | - | 0.82 (0.43, 1.55) | 0.81 (0.43, 1.53) | 0.81 (0.43, 1.53) |
| Benign or non-atypical proliferative breast changes | | | | | |
| Fibroadenoma | 191 (11.9%) | 14 (7.3%) | 0.98 (0.56, 1.70) | 0.96 (0.55, 1.67) | 0.96 (0.55, 1.68) |
| Calcification | 510 (31.8%) | 32 (6.3%) | 0.82 (0.55, 1.23) | 0.80 (0.53, 1.20) | 0.82 (0.54, 1.23) |
| Cyst | 257 (16.0%) | 14 (5.4%) | 0.69 (0.39, 1.20) | 0.68 (0.39, 1.18) | 0.67 (0.38, 1.17) |
| Apocrine metaplasia | 215 (13.4%) | 10 (4.7%) | 0.56 (0.29, 1.07) | 0.57 (0.30, 1.08) | 0.53 (0.28, 1.03) |
| Columnar cell change | 180 (11.2%) | 9 (5.0%) | 0.73 (0.37, 1.44) | 0.76 (0.38, 1.50) | 0.77 (0.39, 1.52) |
| Usual ductal hyperplasia (UDH) | 211 (13.2%) | 18 (8.5%) | 1.26 (0.77, 2.08) | 1.25 (0.76, 2.07) | 1.25 (0.76, 2.07) |
| Sclerosing adenosis | 189 (11.8%) | 14 (7.4%) | 1.07 (0.61, 1.87) | 1.09 (0.63, 1.90) | 1.12 (0.64, 1.96) |
| Fibrocystic change | 491 (30.6%) | 38 (7.7%) | 1.12 (0.76, 1.64) | 1.15 (0.79, 1.69) | 1.18 (0.80, 1.74) |
| Any (binary) | 914 (57.0%) | 65 (7.1%) | 0.86 (0.61, 1.22) | 0.86 (0.61, 1.22) | 0.86 (0.60, 1.21) |
| Any (continuous) | - | - | 0.96 (0.86, 1.07) | 0.96 (0.86, 1.07) | 0.96 (0.86, 1.08) |
| Early neoplastic, papillary and complex sclerosing lesions | | | | | |
| Ductal carcinoma in situ (DCIS) | 1073 (66.9%) | 84 (7.8%) | 0.99 (0.69, 1.43) | 0.94 (0.65, 1.37) | 0.97 (0.67, 1.40) |
| Atypical ductal hyperplasia (ADH) | 93 (5.8%) | 6 (6.5%) | 0.91 (0.40, 2.06) | 0.89 (0.39, 2.03) | 0.91 (0.40, 2.09) |
| Flat epithelial atypia (FEA) | 49 (3.1%) | 1 (2.0%) | 0.35 (0.05, 2.54) | 0.38 (0.05, 2.75) | 0.39 (0.05, 2.83) |
| Radial scar or complex sclerosing lesion | 74 (4.6%) | 8 (10.8%) | 1.39 (0.68, 2.84) | 1.57 (0.76, 3.24) | 1.63 (0.79, 3.36) |
| Intraductal papilloma | 129 (8.0%) | 16 (12.4%) | **1.69 (1.00, 2.86)** | **1.71 (1.01, 2.90)** | **1.75 (1.03, 2.98)** |
| Any (binary) | 1138 (70.9%) | 94 (8.3%) | 1.21 (0.81, 1.79) | 1.17 (0.78, 1.74) | 1.20 (0.81, 1.79) |
| Any (continuous) | - | - | 1.08 (0.86, 1.37) | 1.08 (0.85, 1.37) | 1.10 (0.87, 1.40) |

**Supplementary Table 9.** Association between coexisting features and 10-year overall survival in 1,634 breast cancer cases with reports from excision procedures diagnosed with Stage II breast cancer. Hazard ratios (HRs) with 95% confidence intervals (CIs) are presented from Cox proportional hazards models evaluating the impact of coexisting breast features on 10-year overall survival among breast cancer patients. All tests were two-sided. Statistically significant associations ($p < 0.05$) are indicated in bold. Multiple comparisons were controlled using the Benjamini–Hochberg procedure (false discovery rate adjustment); associations remaining significant after correction are denoted by *. a) Adjusted for age at diagnosis and year of diagnosis. b) Model one further adjusted for menstruation status, ethnicity, family history of cancer and parity. c) Model two further adjusted for tumour characteristics: subtype.

| | Number of patients | Number of events | Model 1[a] HR (95% CI) | Model 2[b] HR (95% CI) | Model 3[c] HR (95% CI) |
|---|---|---|---|---|---|
| Lobular neoplasia | | | | | |
| Lobular carcinoma in situ (LCIS) | 94 (5.8%) | 14 (14.9%) | 1.04 (0.60, 1.78) | 1.08 (0.63, 1.87) | 1.15 (0.66, 2.00) |
| Atypical lobular hyperplasia (ALH) | 20 (1.2%) | 5 (25.0%) | 1.92 (0.79, 4.65) | 2.13 (0.86, 5.27) | 2.18 (0.88, 5.41) |
| Any (binary) | 116 (7.1%) | 14 (14.9%) | 1.15 (0.70, 1.89) | 1.22 (0.74, 2.00) | 1.30 (0.78, 2.15) |
| Any (continuous) | - | - | 1.16 (0.75, 1.78) | 1.22 (0.79, 1.87) | 1.28 (0.83, 1.98) |
| Benign or non-atypical proliferative breast changes | | | | | |
| Fibroadenoma | 192 (11.8%) | 26 (13.5%) | 0.94 (0.62, 1.41) | 0.93 (0.62, 1.41) | 0.97 (0.64, 1.46) |
| Calcification | 351 (21.5%) | 47 (13.4%) | 0.92 (0.67, 1.27) | 0.91 (0.66, 1.26) | 0.93 (0.68, 1.29) |
| Cyst | 255 (15.6%) | 33 (12.9%) | 0.96 (0.67, 1.39) | 0.98 (0.67, 1.42) | 0.99 (0.68, 1.44) |
| Apocrine metaplasia | 157 (9.6%) | 22 (14.0%) | 1.08 (0.70, 1.67) | 1.06 (0.68, 1.65) | 1.06 (0.68, 1.65) |
| Columnar cell change | 120 (7.3%) | 17 (14.2%) | 1.13 (0.69, 1.86) | 1.17 (0.71, 1.92) | 1.22 (0.74, 2.00) |
| Usual ductal hyperplasia (UDH) | 160 (9.8%) | 15 (9.4%) | 0.63 (0.37, 1.07) | 0.62 (0.37, 1.05) | 0.64 (0.38, 1.08) |
| Sclerosing adenosis | 135 (8.3%) | 21 (15.6%) | 1.25 (0.80, 1.96) | 1.24 (0.79, 1.95) | 1.32 (0.84, 2.09) |
| Fibrocystic change | 399 (24.4%) | 51 (12.8%) | 0.91 (0.66, 1.23) | 0.91 (0.66, 1.24) | 0.91 (0.67, 1.24) |
| Any (binary) | 803 (49.1%) | 109 (13.6%) | 0.89 (0.69, 1.15) | 0.90 (0.69, 1.16) | 0.93 (0.72, 1.21) |
| Any (continuous) | - | - | 0.98 (0.89, 1.07) | 0.98 (0.89, 1.07) | 0.99 (0.90, 1.08) |
| Early neoplastic, papillary and complex sclerosing lesions | | | | | |
| Ductal carcinoma in situ (DCIS) | 928 (56.8%) | 137 (14.8%) | 1.02 (0.79, 1.32) | 1.02 (0.78, 1.32) | 1.10 (0.84, 1.44) |
| Atypical ductal hyperplasia (ADH) | 62 (3.8%) | 7 (11.3%) | 0.77 (0.36, 1.63) | 0.81 (0.38, 1.72) | 0.85 (0.40, 1.82) |
| Flat epithelial atypia (FEA) | 44 (2.7%) | 2 (4.5%) | 0.32 (0.08, 1.30) | 0.35 (0.09, 1.42) | 0.39 (0.09, 1.57) |
| Radial scar or complex sclerosing lesion | 45 (2.8%) | 5 (11.1%) | 0.99 (0.41, 2.39) | 0.95 (0.39, 2.31) | 1.00 (0.41, 2.42) |
| Intraductal papilloma | 71 (4.3%) | 8 (11.3%) | 0.67 (0.33, 1.37) | 0.68 (0.34, 1.38) | 0.71 (0.35, 1.44) |
| Any (binary) | 979 (59.9%) | 143 (14.6%) | 1.00 (0.77, 1.29) | 1.00 (0.77, 1.30) | 1.08 (0.83, 1.42) |
| Any (continuous) | - | - | 0.92 (0.76, 1.12) | 0.92 (0.76, 1.13) | 0.98 (0.80, 1.20) |

**Supplementary Table 10.** Association between coexisting features and 10-year overall survival in 750 breast cancer cases with reports from excision procedures diagnosed with Stage III breast cancer. Hazard ratios (HRs) with 95% confidence intervals (CIs) are presented from Cox proportional hazards models evaluating the impact of coexisting breast features on 10-year overall survival among breast cancer patients. All tests were two-sided. Statistically significant associations ($p < 0.05$) are indicated in bold. Multiple comparisons were controlled using the Benjamini–Hochberg procedure (false discovery rate adjustment); associations remaining significant after correction are denoted by *. a) Adjusted for age at diagnosis and year of diagnosis. b) Model one further adjusted for menstruation status, ethnicity, family history of cancer and parity. c) Model two further adjusted for tumour characteristics: subtype.

| | Number of patients | Number of events | Model 1[a] HR (95% CI) | Model 2[b] HR (95% CI) | Model 3[c] HR (95% CI) |
|---|---|---|---|---|---|
| Lobular neoplasia | | | | | |
| Lobular carcinoma in situ (LCIS) | 43 (5.7%) | 16 (37.2%) | 1.19 (0.72, 1.98) | 1.21 (0.73, 2.02) | 1.28 (0.76, 2.14) |
| Atypical lobular hyperplasia (ALH) | 8 (1.1%) | 1 (12.5%) | 0.36 (0.05, 2.60) | 0.32 (0.04, 2.30) | 0.39 (0.05, 2.86) |
| Any (binary) | 47 (6.3%) | 16 (34.0%) | 1.09 (0.66, 1.82) | 1.09 (0.65, 1.82) | 1.16 (0.69, 1.95) |
| Any (continuous) | - | - | 1.04 (0.67, 1.63) | 1.03 (0.66, 1.62) | 1.11 (0.70, 1.75) |
| Benign or non-atypical proliferative breast changes | | | | | |
| Fibroadenoma | 108 (14.4%) | 27 (25.0%) | 0.71 (0.48, 1.06) | 0.74 (0.49, 1.11) | 0.83 (0.55, 1.25) |
| Calcification | 159 (21.2%) | 37 (23.3%) | **0.64 (0.45, 0.90)** | **0.64 (0.45, 0.91)** | **0.66 (0.46, 0.94)** |
| Cyst | 114 (15.2%) | 27 (23.7%) | 0.70 (0.47, 1.04) | 0.72 (0.48, 1.08) | 0.69 (0.46, 1.04) |
| Apocrine metaplasia | 51 (6.8%) | 16 (31.4%) | 0.96 (0.58, 1.60) | 0.99 (0.59, 1.64) | 1.03 (0.62, 1.73) |
| Columnar cell change | 48 (6.4%) | 10 (20.8%) | 0.60 (0.32, 1.13) | 0.66 (0.35, 1.26) | 0.63 (0.33, 1.19) |
| Usual ductal hyperplasia (UDH) | 42 (5.6%) | 12 (28.6%) | 0.93 (0.52, 1.66) | 0.90 (0.50, 1.62) | 0.95 (0.53, 1.71) |
| Sclerosing adenosis | 49 (6.5%) | 11 (22.4%) | 0.72 (0.39, 1.33) | 0.75 (0.41, 1.38) | 0.77 (0.42, 1.41) |
| Fibrocystic change | 152 (20.3%) | 39 (25.7%) | 0.79 (0.56, 1.11) | 0.81 (0.57, 1.14) | 0.83 (0.59, 1.17) |
| Any (binary) | 350 (46.7%) | 101 (28.9%) | **0.77 (0.60, 0.99)** | 0.78 (0.60, 1.01) | 0.80 (0.62, 1.04) |
| Any (continuous) | - | - | **0.87 (0.78, 0.97)** | **0.88 (0.79, 0.98)** | **0.89 (0.80, 0.99)** |
| Early neoplastic, papillary and complex sclerosing lesions | | | | | |
| Ductal carcinoma in situ (DCIS) | 448 (59.7%) | 146 (32.6%) | 0.89 (0.69, 1.14) | 0.92 (0.72, 1.18) | 1.01 (0.78, 1.31) |
| Atypical ductal hyperplasia (ADH) | 15 (2.0%) | 1 (6.7%) | 0.16 (0.02, 1.14) | 0.16 (0.02, 1.15) | 0.18 (0.03, 1.31) |
| Flat epithelial atypia (FEA) | 15 (2.0%) | 5 (33.3%) | 0.97 (0.40, 2.36) | 1.07 (0.43, 2.64) | 1.17 (0.47, 2.92) |
| Radial scar or complex sclerosing lesion | 29 (3.9%) | 4 (13.8%) | **0.34 (0.13, 0.91)** | **0.30 (0.11, 0.82)** | **0.33 (0.12, 0.89)** |
| Intraductal papilloma | 40 (5.3%) | 17 (42.5%) | 1.53 (0.93, 2.51) | 1.62 (0.98, 2.70) | **1.74 (1.05, 2.90)** |
| Any (binary) | 471 (62.8%) | 156 (33.1%) | 0.92 (0.72, 1.19) | 0.95 (0.73, 1.22) | 1.05 (0.81, 1.37) |
| Any (continuous) | - | - | 0.88 (0.73, 1.07) | 0.90 (0.74, 1.08) | 0.97 (0.80, 1.17) |

**Supplementary Table 11.** Association between tumor stage and presence/number of breast features, using 3,988 breast cancer cases with reports from excision procedures. All tests were two-sided. No multiple comparison adjustment was applied as analyses were pre-specified and limited in number.

| Model type | Predictor (ref Stage I) | Estimate | 95% confidence interval | p-value |
|---|---|---|---|---|
| Logistic (binary) | Stage II | 0.597 | (0.501, 0.710) | 6.35E-09 |
| | Stage III | 0.681 | (0.548, 0.847) | 5.28E-04 |
| Poisson (count) | Stage II | 0.779 | (0.743, 0.817) | 8.52E-25 |
| | Stage III | 0.739 | (0.694, 0.786) | 2.61E-21 |

**Supplementary Figure 1.** Flowchart of how analytical datasets were derived.

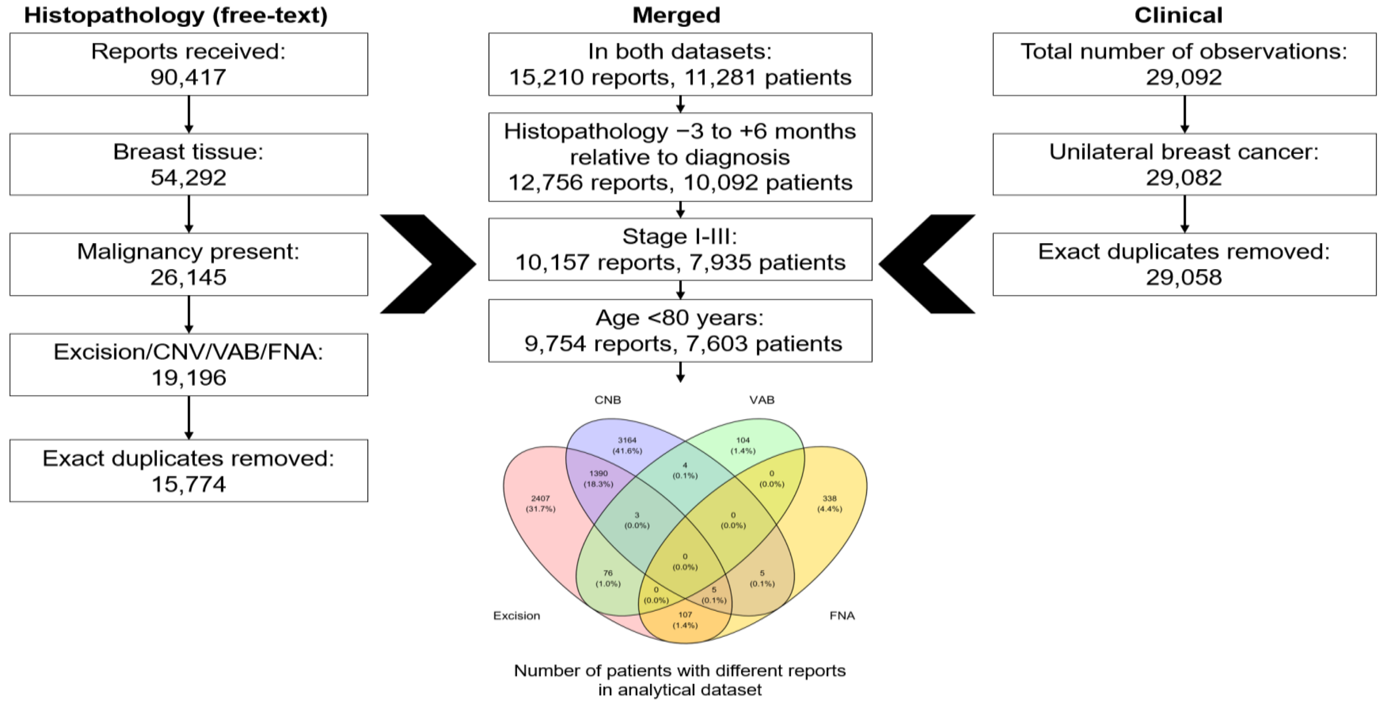

**Supplementary Figure 2.** Distribution of procedure date relative to diagnosis date.

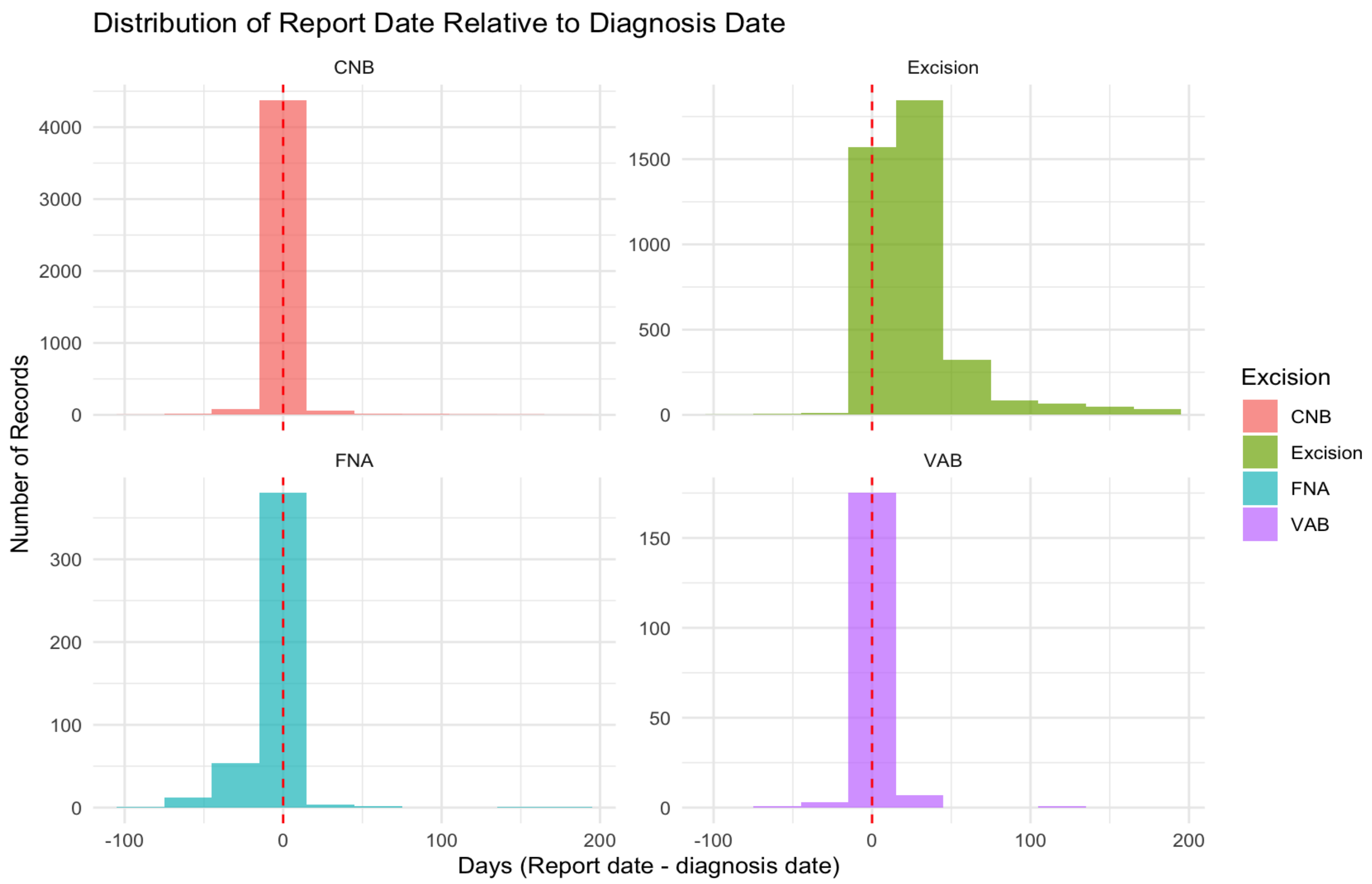

**Supplementary Figure 3.** Cluster membership of breast features based on Pearson correlation hierarchical clustering for 3,988 records from excisions.

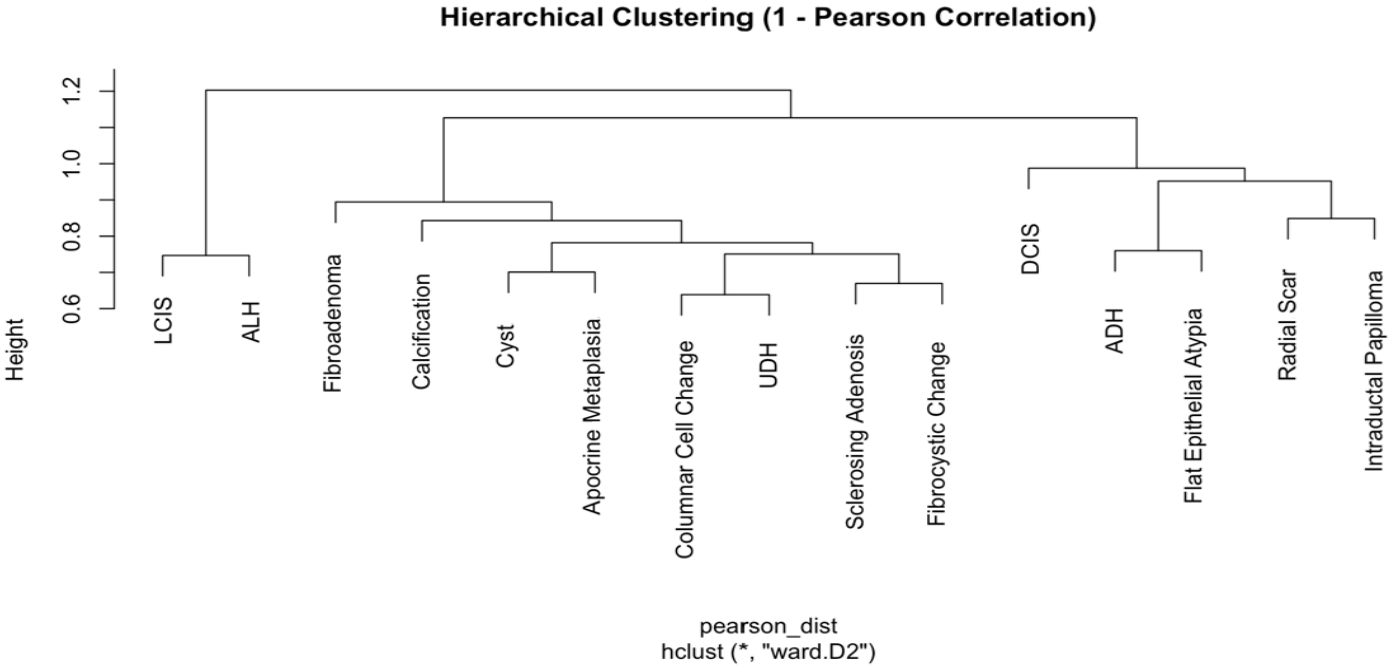
